# A Genome-wide Association Study Identifies Novel Genetic Variants Associated with Knee Pain in the UK Biobank (N = 441,757)

**DOI:** 10.1101/2024.09.16.24313726

**Authors:** Yiwen Tao, Qi Pan, Tengda Cai, Luning Yang, Mainul Haque, Tania Dottorini, Weihua Meng

**Affiliations:** Nottingham Ningbo China Beacons of Excellence Research and Innovation Institute, University of Nottingham Ningbo China, Ningbo, China, 315100; School of Mathematical Sciences, University of Nottingham Ningbo China, Ningbo, China, 315100; School of Veterinary Medicine and Science, University of Nottingham, Nottingham, UK, LE12 5RD; Division of Population Health and Genomics, Ninewells Hospital and Medical School, University of Dundee, Dundee, UK, DD2 4BF; Center for Public Health, Faculty of Medicine, Health and Life Sciences, School of Medicine, Dentistry and Biomedical Sciences, Queen’s University Belfast, Belfast, UK, BT12 6BA

**Keywords:** knee pain, genome-wide association study, genetic correlations, phenome-wide association analysis, transcriptome-wide association study, Mendelian randomization

## Abstract

Knee pain is a widespread musculoskeletal condition affecting millions globally, with significant socio-economic implications. This study endeavors to identify genetic variants associated with knee pain through a comprehensive genome-wide association study (GWAS) using data from 441,757 individuals in the UK Biobank. The primary GWAS identified ten significant loci, including eight novel loci, with the most significant single nucleotide polymorphism (SNP) being rs143384 near the *GDF5* gene on chromosome 20 (*p* = 4.68 × 10^-19^). In the replication study, seven loci (rs143384, rs919642, rs55760279, rs56076919, rs3892354, rs687878, rs368636424) were found to be significant in the FinnGen cohort. Further, sex-specific analyses revealed distinct genetic associations, identifying three loci (rs143384 with *p* = 1.70×10^-15^, rs56076919 with *p* = 1.60×10^-9^, rs919642 with *p* = 1.45×10^-8^) in females and four loci ( rs2899611 with *p* = 2.77 × 10^-11^, rs891720 with *p* = 5.55 × 10^-11^, rs2742313 with *p* = 4.19 × 10^-9^, rs2019689 with *p* = 6.51 × 10^-9^) in males. The phenome-wide association analysis and Mendelian randomization analysis revealed significant links between several phenotypes and knee pain such as leg pain on walking. These findings enhance our understanding of the genetic factors of knee pain, offering potential pathways for therapeutic interventions and personalized medical strategies.

## Introduction

Knee pain is a prevalent musculoskeletal complaint that significantly impacts the quality of life and functional ability of millions worldwide (Jinks et al. 2008; Neogi 2013). It ranks among the most common musculoskeletal complaints, particularly in older adults, and is a leading cause of functional impairment and healthcare burden worldwide (Cross et al. 2014). Knee pain can arise from various etiologies, including osteoarthritis (OA), injury, inflammation, and other degenerative joint diseases (Dulay et al. 2015; Huang et al.). Epidemiological studies highlight the considerable prevalence of knee pain, with a survey in British hospitals showing that the 12-month period occurrence of knee pain in the entire population aged 50 and over is 46.8% (C et al. 2004). However, few studies have focused specifically on the genetic factors contributing to knee pain. In most early studies, standard outcomes have been evaluated by radiology for OA, which may be due to the benefit of an objective definition of the condition (Miranda et al. 2002). Knee pain is often caused by other conditions rather than OA, therefore, genetic factors may also differ.

Multiple factors contribute to the occurrence and progression of knee pain, including age, sex, obesity, physical activity, and occupational hazards. A cross-sectional survey showed that 8.1% of men and 23.5% of women in the United States aged 60 years and older reported knee pain, with men reporting knee pain less frequently than women of the same age (Andersen et al. 1999). Obesity is another significant risk factor. A survey of hospitals in England showed that knee pain prevalence ranged from 23% in the normal-weight category to 31% in the obese group, showing a moderate association between obesity and subsequent knee pain (Jinks et al. 2006). Physical activities that involve repetitive stress or injury to the knee, as well as occupations that require prolonged kneeling or heavy lifting, also elevate the risk of knee pain (Cooper et al. 1994).

Genetic predisposition plays a pivotal role in the susceptibility to knee pain, with accumulating evidence underscoring the importance of hereditary factors in its development. A previous genome-wide association study (GWAS) has identified genes *GDF5* and *COL27A1* that are associated with knee pain, highlighting the heritable nature of this condition (Meng et al. 2019). Additionally, GWAS have identified genes associated with knee OA, including *GNL3*, *ASTN2*, *FILIP1*, *SENP6*, *CHST11, FTO and BTNL2* (Nakajima et al. 2010), although the relationship between these genes and knee pain specifically has not been fully clarified.

This study aims to identify new genetic variants associated with knee pain by conducting a GWAS utilizing data from the UK Biobank using new definitions of a larger control group. The findings were replicated and compared to the FinnGen Biobank cohort (Kurki et al. 2023). We also conducted novel sex-specific GWAS to investigate possible genetic variations linked with males or females. Two-sample Mendelian Randomization (MR) was further employed to investigate potential causal relationships between knee pain and various phenotypes.

## Methods

### Cohort Information

The UK Biobank is an extensive cohort designed to support research into various health conditions in more than 500,000 people aged 40-69. It includes a range of information about their health and genetic data from the beginning of recruitment in 2006 until 2010 in the UK. Participants provided informed consent to complete comprehensive questionnaires and to donate biological samples such as blood, saliva and urine (further details accessible at www.ukbiobank.ac.uk). Ethical approval for this study was granted by the National Research Ethics Service and the National Health Service (reference 11/NW/0382). The genetic and associated data from the UK Biobank are accessible for authorized research programs investigating a vast variety of illnesses.

DNA extraction and subsequent quality control (QC) processes were standardized before the data release to the UK Biobank, detailed in the https://biobank.ctsu.ox.ac.uk/crystal/ukb/docs/genotyping_sample_workflow.pdf. The Wellcome Trust Centre for Human Genetics at Oxford University ensured the reliability of the genotyping results. QC measures included identifying underperforming markers, evaluating sample correlations, and adjusting for batch effects, according to the information found at http://biobank.ctsu.ox.ac.uk/crystal/refer.cgi?id=155580.

### Definitions of Case and Control

The UK Biobank questionnaire included a specific question about pain, used to define the case group and the control group in the study: “In the last month have you experienced any of the following that interfered with your usual activities? (You can select more than one answer)” Response options included seven different body sites (head, face, neck or shoulder, back, stomach or abdomen, hip, and knee), “all over the body,” “none of the above,” and “prefer not to answer” (UK Biobank Field ID: 6159). Participants could select multiple answers. Cases for knee pain were those who answered “yes” to “Knee pain”. Controls were defined as those who did not report knee pain and did not choose “Prefer not to say.” To minimize population stratification, only data from white British participants (Field ID: 21000) were included.

### Design and replication of the GWAS

The primary goal of this GWAS was to identify genetic variants linked to knee pain through GWAS. Given the potential differences in genetic architecture between sexes, additional sex-specific GWAS analyses were conducted to uncover any sex-related genetic differences in knee pain. We employed the publicly accessible summary statistics of knee arthrosis from the FinnGen dataset during the replication phase (Kurki et al. 2023)

### GWAS and Subsequent Statistical Analysis

Genome-wide complex trait analysis (GCTA, v1.94.1) was employed to estimate the genetic contribution of knee pain, which is accessible at https://yanglab.westlake.edu.cn/software/gcta/#Overview (Yang et al. 2011). The fastGWA function in GCTA, which uses a generalized mixed linear model association approach, was utilized for the GWAS association analysis with a sparse genetic relationship matrix. QC measures included exclusion of minor allele frequencies less than 0.5%, single nucleotide polymorphisms (SNPs) with INFO scores less than 0.3 and those failing Hardy-Weinberg equilibrium tests (*p* < 1 × 10^-6^). Mitochondrial SNPs along with SNPs on sex chromosomes were also eliminated. The association tests had adjustments for age, sex, BMI and eight main components. Data from white British participants were processed using R v4.2.2 to identify cases and controls. Chi-square tests and independent t-tests were used to evaluate differences in sex frequency and other covariates between cases and controls, with a significance threshold of *p* < 0.05. A genome-wide significance criterion of *p* < 5×10^-8^ was used. Additionally, GCTA was used to estimate narrow-sense heritability.

### Functional Analysis

Functional Mapping and Annotation (FUMA) of GWAS results was performed using the SNP2GENE function, which offers thorough functional annotation for SNPs in genomic areas highlighted by lead SNPs (Watanabe et al. 2017). FUMA default parameters were used, including significant SNP thresholds (p < 5×10^-8^) and linkage disequilibrium blocks (r^2^ > 0.6), as well as a minimum minor allele frequency greater than 0.01. The maximum distance between linkage disequilibrium blocks to merge into a locus was set at 250 kb, with the 1000G Phase3 EUR population used as the reference. Regional visualizations were generated using Locus Zoom (Pruim et al. 2010).

FUMA facilitates three primary analytical approaches including gene-based association analysis, gene-set analysis, and tissue expression analysis. For gene-based and gene-set analyses, FUMA integrates MAGMA (v1.0619) as a key tool. In gene-based association analysis, the summary statistics of SNPs are aggregated at the gene level. Statistical data from SNPs at each locus are combined to assess the association between entire genes and the phenotype under study. Specifically, SNPs located within genes are mapped to 19,023 protein-coding genes, and significance is determined with a threshold of *p* = 0.05/19,023 = 2.60 × 10^-6^. This integration allows us to evaluate the association score between each gene and the studied phenotype.

In gene-set analysis, FUMA conducts a comprehensive examination of collections of genes that share common biological functions or other characteristics. The significance threshold for this analysis is *p* = 0.05/15,485 = 3.23 × 10^-6^.

For tissue expression analysis, FUMA utilizes tissue-specific expression data from the GTEx project to evaluate the expression levels of specific genes across different tissues. We constructed four gene expression heat maps for genes pinpointed through location mapping, using average and normalized mean expression values for each label. The data for these heatmaps were derived from GTEx v8 datasets, encompassing 54 distinct tissue types and 30 general tissue categories. Tissue specificity was assessed using differentially expressed genes (DEGs) predefined for each label from the expression datasets. Enrichment evaluations for DEGs were conducted for both tissue types from GTEx v8, focusing specifically on the enrichment of positional mapping genes without considering the overall distribution of SNP *p* values from the MAGMA tissue expression analysis.

### Integration of eQTL Analysis, Chromatin Architecture, and Positional Mapping

Expression quantitative trait loci (eQTL) are crucial for elucidating the regulatory mechanisms associated with variants discovered through GWAS (Võsa et al. 2021). Cis-eQTLs, in particular, modulate gene expression by interacting with nearby variants within a 1 Mb range, thus directly influencing gene regulation. In eukaryotic cells, the genome is intricately packed within the nucleus, with chromatin serving as the essential structural component. This compact organization is key to establishing a three-dimensional genomic structure necessary for DNA replication, repair, gene transcription, and other fundamental biological processes. In this research, we applied positional mapping using a 10 kb distance threshold. This approach integrates cis-eQTL analysis, chromatin interaction data, and positional mapping to provide a comprehensive understanding of the genomic landscape.

### Genetic Correlation Analysis using LDSC

Linkage disequilibrium score regression (LDSC) is a statistical technique utilized to estimate genetic correlations and heritability from the GWAS summary statistics, detailed at https://github.com/bulik/ldsc (Bulik-Sullivan et al. 2015). LDSC assesses the relationship between various phenotypes by measuring the linkage disequilibrium between pairs of loci. Additionally, LDSC can generate a genome-wide map of genetic correlations, highlighting clusters of loci linked to a certain disease or trait. LDSC was employed to examine the genetic correlation of knee pain between males and females, aiming to identify overall genetic differences between these patient groups. Through Complex-Traits Genetics Virtual Lab (https://genoma.io/), we also analyzed genetic connections of knee pain using 1,396 characteristics from the UK Biobank. This open-source platform consolidates GWAS datasets to perform genetic correlation analyses of complex traits using LDSC. The results were adjusted for multiple testing using the Bonferroni correction method.

### Transcriptome-wide Association Studies (TWAS)

We used Transcriptome-Wide Association Studies (TWAS) (http://gusevlab.org/projects/fusion/) to evaluate how genetic variations influence gene expression in specific tissue(Gusev et al. 2016). TWAS analyzed SNP-gene expression relationships from eQTL studies and SNP-disease connections from GWAS summary data for particular tissues in GTEx v7 panel.

### Phenome-Wide Association Study (PheWAS) Analysis

The Phenome-Wide Association Study (PheWAS) analysis was performed to examine the relationships between significant SNPs and their associated genes across a broad spectrum of traits. This analysis had two main objectives including corroborating GWAS findings by establishing connections with pain-related phenotypes, and uncovering novel associations between genetic variants implicated in knee pain and other phenotypes, with a particular focus on psychiatric traits such as OA, as outlined by the ATLAS platform at https://atlas.ctglab.nl/PheWAS (Watanabe et al. 2019). This investigation utilized a dataset comprising 4,756 GWAS summary statistics available on the GWAS ATLAS platform (Watanabe et al. 2019). Only SNPs with *p* values below 0.05 were included in the analysis, and the Bonferroni correction was applied to adjust for multiple comparisons.

### Mendelian Randomization

This research utilized a two-sample MR approach to examine the potential causal relationship between several phenotypes and knee pain. Based on our genetic correlation analysis (T > 0.7) and PheWAS findings, MR was used to ascertain whether these genetic loci exert direct effects on knee pain, independent of a causal pathway. Genetic associations for the examined phenotypes were sourced from the UK Biobank genetic databases via the IEU Open GWAS platform (https://gwas.mrcieu.ac.uk/). The genetic data on knee pain were sourced from our primary GWAS, which included 101,544 cases and 338,199 controls. We applied Inverse Variance Weighted (IVW) estimation, supplemented by MR Egger, Weighted Median, Simple Mode, and Weighted Mode methods to ensure robustness in our sensitivity analyses. To address potential biases, we conducted heterogeneity assessments using Cochran’s Q test and evaluated horizontal pleiotropy with the MR Egger intercept test. Furthermore, bidirectional MR analyses were performed to explore the possibility of reverse causation.

## Results

### Description of the samples

The initial phase of the UK Biobank study, conducted from 2006 to 2010, involved the participation of 501,708 individuals who completed a pain questionnaire. Among this large cohort, 116,560 individuals reported experiencing ‘knee pain,’ leading to their categorization as ‘cases’ in the study, while 384,358 participants who did not report such specific pain were classified as ‘controls’. Following the refinement of the data, which involved the inclusion of only white-British participants who met the QC criteria, the primary GWAS analysis encompassed 101,544 cases (53,078 males and 48,466 females) and 338,199 controls (185,718 males and 152,481 females). Subsequently, the study entered a secondary phase, which involved a more detailed, sex-stratified GWAS analysis. The QC procedures were consistently applied, resulting in a female cohort of 238,796 samples (split into 53,078 cases and 185,718 controls) and a male cohort of 200,947 samples (48,466 cases and 152,481 controls). An overview of the clinical attributes for both case and control groups is provided in Table 1. Statistically significant discrepancies were observed in variables such as age, sex, and body mass index (BMI) between case and control groups, registering a *p* value of less than 0.001, establishing the differences as statistically substantive.

**Table 1.** Clinical characteristics of knee pain cases and controls in the UK Biobank.

| GWAS Analysis | Covariates | Cases | Controls | *p*-value |
| --- | --- | --- | --- | --- |
| Primary GWAS | Sex (female: male) | 53,078 : 48,466 | 185,718 : 152,481 | <0.001 |
|  | Age (years) | 57.6 (7.72) | 56.6 (8.07) | <0.001 |
|  | BMI (kg/m<sup>3</sup>) | 28.9 (5.28) | 27.0 (4.52) | <0.001 |
| Female-specific GWAS | Age (years) | 57.7 ( 7.51 ) | 56.3 ( 8.00 ) | <0.001 |
|  | BMI (kg/m3) | 28.9 ( 5.80) | 26.5 ( 4.83 ) | <0.001 |
| Male-specific GWAS | Age (years) | 57.6 ( 7.93 ) | 56.9 ( 8.13 ) | <0.001 |
|  | BMI (kg/m3) | 29.0 ( 4.65 ) | 27.5 ( 4.05 ) | <0.001 |

Total number of females: 238,796; Total number of males: 200,947.
A $\chi^2$ test was used to test the difference of gender frequency between cases and controls and an independent t test was used for other covariates.
Continuous covariates were presented as mean (standard deviation)
BMI represents body mass index.

### GWAS Results

During the primary stage of the GWAS investigation, we identified ten significant GWAS loci that exhibited genome-wide significance of *p* values < 5 × 10^-8^, as depicted in Figure 1. Notably, eight of the identified loci represent new discoveries. In-depth information regarding the top SNPs within each of these loci can be found in Table 2, while a comprehensive list of all SNPs in this GWAS is available in Supplementary Table 1.

**Fig. 1.**
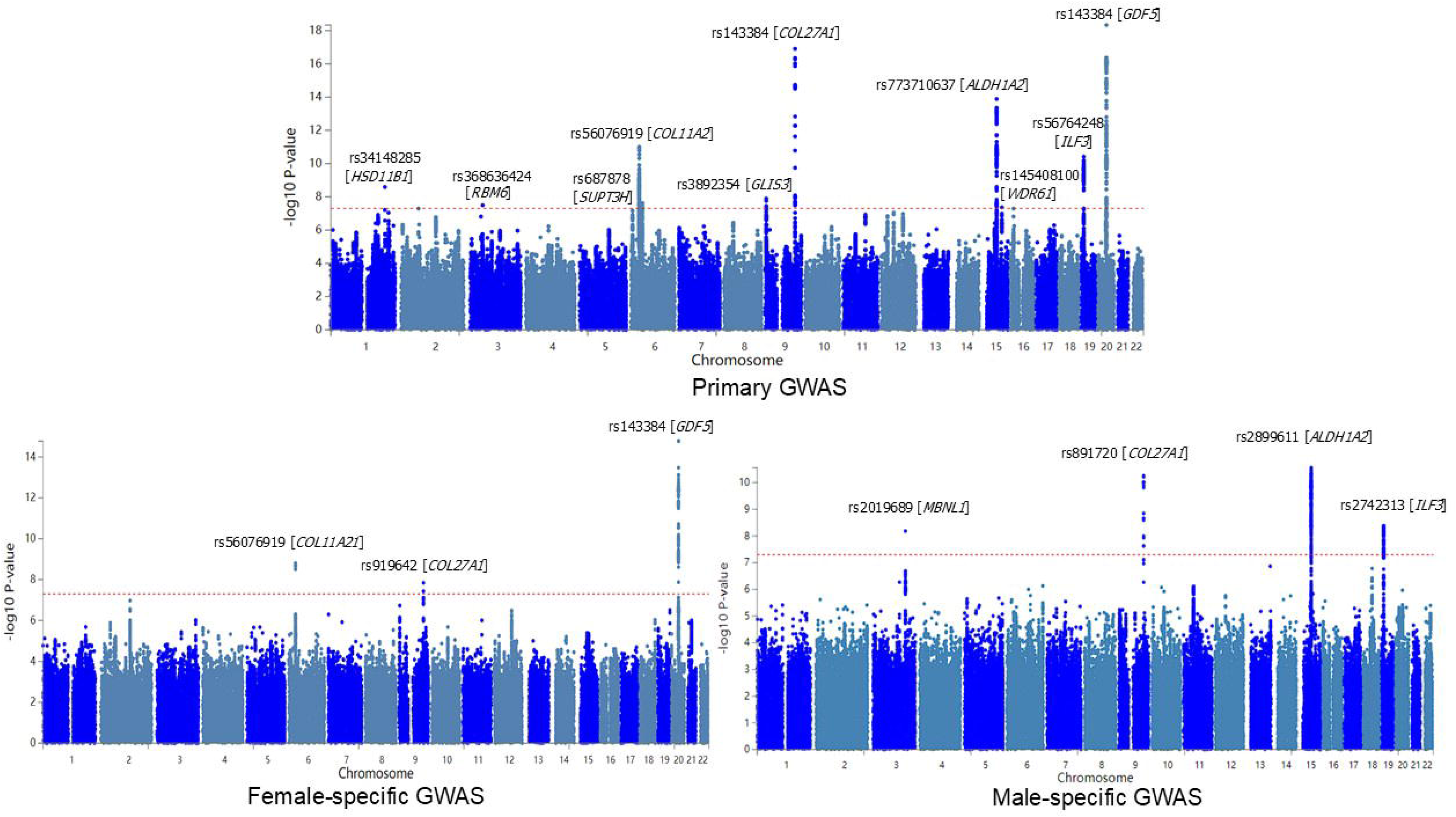
The Manhattan plot of the primary GWAS analysis on knee pain (N = 441,757), the female-specific GWAS analysis (N = 238,796) and the male-specific GWAS analysis (N= 200,947) The dashed red line indicates the cut-off *p* value of 5 × 10^−8^

**Table 2.**
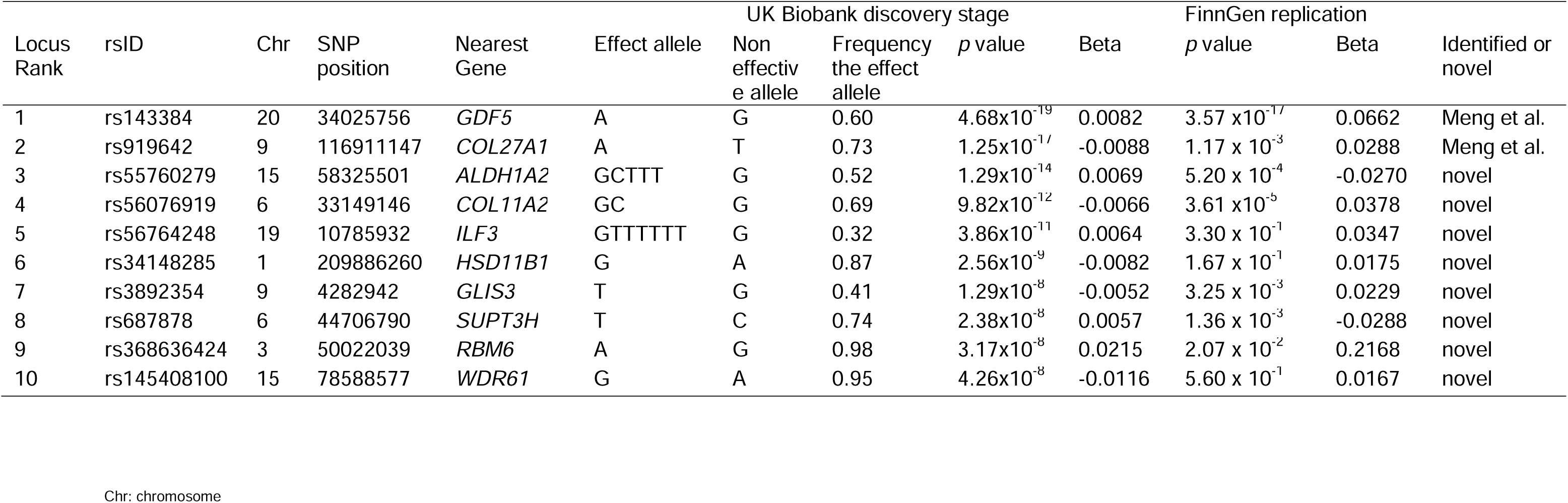
The top SNPs within ten loci identified by the GWAS on knee pain.

In the primary GWAS, the strongest association was observed within the SNP cluster near the growth differentiation factor 5 (*GDF5*) gene on chromosome 12, with rs143384 demonstrating a remarkably low *p* value of 4.68×10^-19^ (regional plot is provided in Figure 2a). The second significant correlations were uncovered in the collagen type XXVII alpha 1 chain (*COL27A1*) gene on chromosome 9, with corresponding *p* values of 1.25×10^-17^ for rs919642 (regional plot is provided in Figure 2b). Among the eight newly identified loci, rs55760279 in the aldehyde dehydrogenase 1 family member A2 (*ALDH1A2*) gene with a *p* value of 1.29×10^-14^ was the most strongly associated (regional plot is provided in Figure 3). The other loci include those with leading SNPs: rs56076919 on chromosome 6, rs56764248 on chromosome 19, rs34148285 on chromosome 1, rs3892354 on chromosome 9, rs687878 on chromosome 6, rs368636424 on chromosome 3, and rs145408100 on chromosome 15. The detailed regional plots for these loci are available in Supplementary Figure 1, while the Q–Q plot of the primary GWAS is shown in the Supplementary Figure 2. The SNP-based heritability for knee pain was estimated at 0.042, with a standard error of 0.0015.

**Fig. 2.**
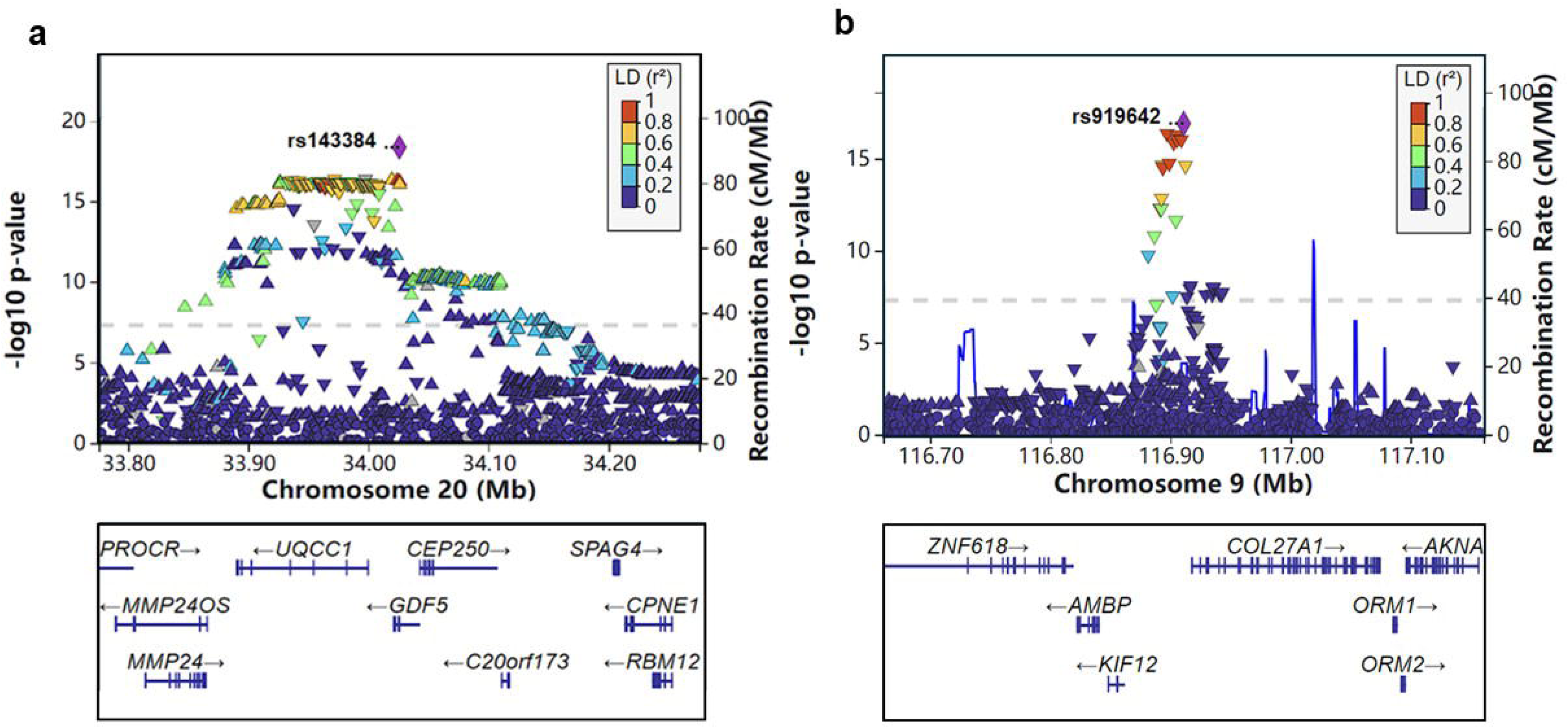
The regional plots of loci in the *GDF5* and *COL27A1* regions

**Fig. 3.**
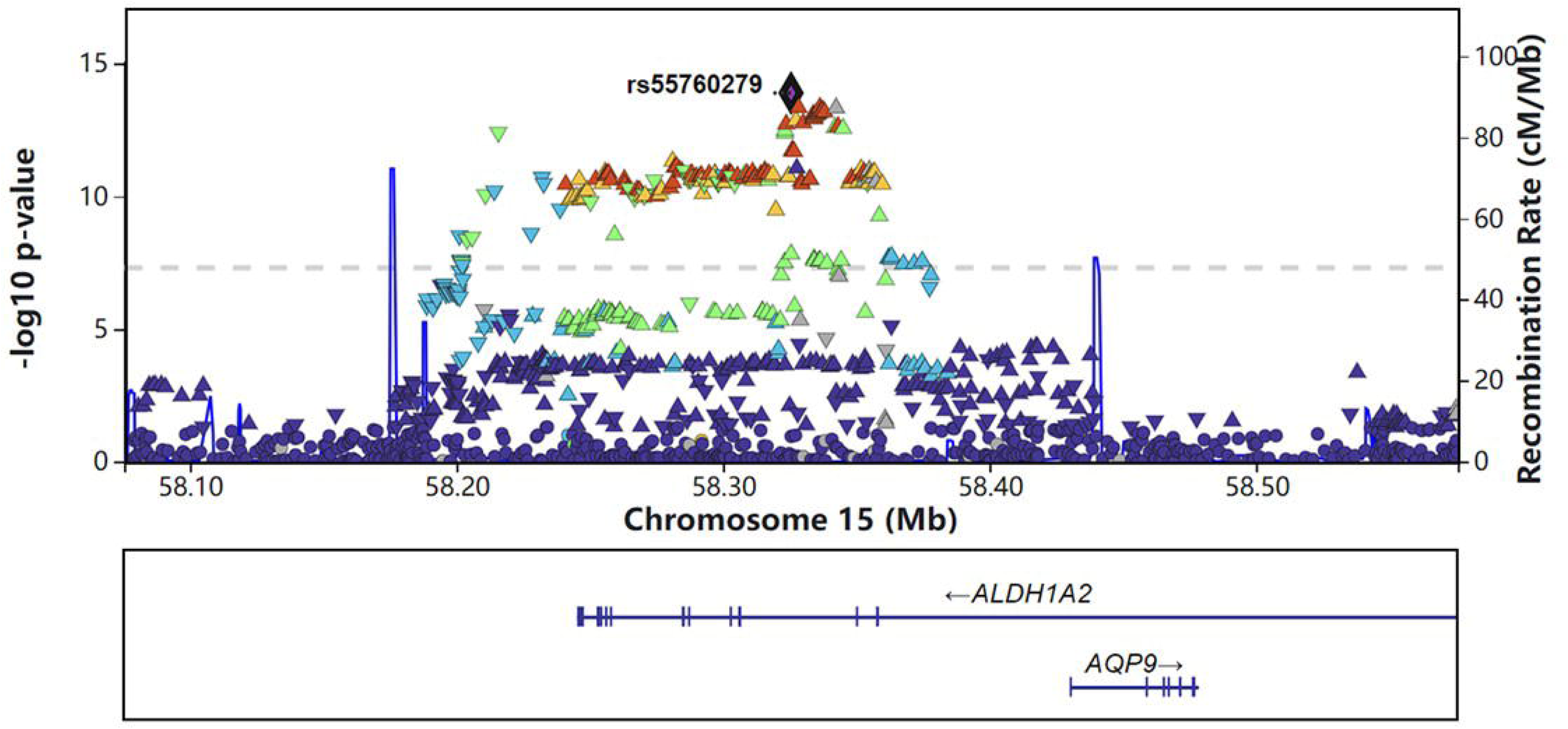
The regional plot of locus in the *ALDH1A2* region

The *p* values for the associations of the ten independent and significant SNPs identified in our primary GWAS were examined in external datasets to assess replication. We observed significant replication (p < 0.05) for seven loci, including those on chromosome 20 (rs143384, *p* = 3.57 × 10^-1^), chromosome 9 (rs919642, *p* = 1.17 × 10^-3^), chromosome 15 (rs55760279, *p* = 5.20 × 10^-4^), chromosome 6 (rs687878, *p* = 1.36 × 10^-3^), chromosome 3 (rs368636424, *p* = 2.07 × 10^-2^), chromosome 6 (rs56076919, *p* = 3.61 × 10^-5^), and chromosome 9 (rs3892354, *p* = 3.25 × 10^-3^). While loci on chromosomes 19 (rs56764248, *p* = 3.30 × 10^-1^), 1 (rs34148285, *p* = 1.67 × 10^-1^) and 15 (rs145408100, *p* = 5.60 × 10^-1^) did not reach the replication threshold (Table 2).

In the subsequent sex-stratified GWAS, the female-specific evaluation revealed three significant loci related to knee pain. These loci were identical to the three most significant loci were consistent with the primary GWAS findings, with lead SNPs rs143384, rs56076919 and rs919642. The male-specific GWAS revealed four significant loci associated with knee pain. The three most significant loci were the same as those identified in the primary GWAS, with lead SNP rs2899611, rs891720 and rs2742313. The novel locus was identified near the muscleblind like splicing regulator 1 (*MBNL1*) gene on chromosome 3, with a *p* value of 6.51×10^-9^ for rs2019689. Further details of these findings are presented in Table 3, with the corresponding Manhattan plots displayed in Figure 1.

**Table 3.** The top SNPs within three loci for the female-specific GWAS and four loci for the male-specific GWAS on knee pain.

| Secondary GWAS | Locus Rank | rsID | Chr | SNP position | Nearest Gene | Effect allele | Non effective allele | Frequency the effect allele | p value | Beta |
| --- | --- | --- | --- | --- | --- | --- | --- | --- | --- | --- |
| Female-specific GWAS | 1 | rs143384 | 20 | 34025756 | *GDF5* | A | G | 0.60 | $1.70 \times 10^{-15}$ | 0.0097 |
| | 2 | rs56076919 | 6 | 33149146 | *COL11A2* | GC | G | 0.69 | $1.60 \times 10^{-9}$ | -0.0078 |
| | 3 | rs919642 | 9 | 116911147 | *COL27A1* | A | T | 0.73 | $1.45 \times 10^{-8}$ | -0.0078 |
| Male-specific GWAS | 1 | rs2899611 | 15 | 58327347 | *ALDH1A2* | T | G | 0.49 | $2.77 \times 10^{-11}$ | 0.0090 |
| | 2 | rs891720 | 9 | 116897213 | *COL27A1* | C | A | 0.73 | $5.55 \times 10^{-11}$ | -0.01006 |
| | 3 | rs2742313 | 19 | 10799750 | *ILF3* | T | C | 0.37 | $4.19 \times 10^{-9}$ | 0.0082 |
| | 4 | rs2019689 | 3 | 152177708 | *MBNL1/*<br>*RP11-362A9.3* | T | C | 0.80 | $6.51 \times 10^{-9}$ | -0.0104 |

Chr: chromosome

### Gene, Gene-set, and Tissue-Specific Expression Analysis Using FUMA

In our comprehensive gene analysis derived from primary GWAS, *GDF5* emerged as the most significantly associated gene. Beyond *GDF5,* our exploration also uncovered significant ties with 18 other genes - namely *SMG7*, *RING1*, *COL11A2*, *HLA-DPB1*, *GLIS3*, *SOX5*, *DERA*, *ALDH1A2*, *USP8*, *UQCC1*, *CEP250*, *ILF3*, *GDF5OS*, *AC011475.1*, *C20orf173*, *SLC44A2*, *QTRT1* and *ERGIC3*, shown in Supplementary Figure 3. Each of these demonstrated a significant relationship with knee pain and yielded *p* values less than 2.60 × 10^-6^, in compliance with the thresholds established post multiple testing adjustments via Bonferroni correction (0.05/19,203).

In addition to dissecting individual genes, we broadened our approach to incorporate a comprehensive analysis of 15,485 gene sets. Within this extensive group, the gene set ‘NIKOLSKY_BREAST_CANCER_20Q11_AMPLICON’ exhibited a prominent association, as represented by a *p* value of 2.37 × 10^-10^. The top ten gene sets from this assessment were furnished in Supplementary Table 2.

In terms of tissue expression analysis, we encompassed a diversified spectrum of 30 general and 53 specific tissue types. However, no significant results under investigation was found from these tissue types. Refer to Supplementary Figure 4 for a more detailed visual representation and elucidation of these findings.

In the gene expression heatmaps as shown in Supplementary Figure 5, genes *ECSIT*, *DNM2*, *AHCY*, *ILF3*, *RXRB*, *CPNE1*, *RBM6*, *SLC39A7*, *RING1*, *ROMO1*, *TRPC4AP*, *ERGIC3*, *RBM39*, *C20orf24*, *EIF6*, *SCAND1* demonstrated high expression across all 54 tissue types, whlie genes *QTRT1*, *RSL24D1* were highly expressed in all tissue types except blood and *SLC44A2* only show weak expression in liver. The average of normalized expression allows comparison of gene expression across labels within a gene. Genes *C17ORF112*, *MNS1* and *CNBD2* were most significantly expressed in testis.

### Integration of eQTL Analysis, Chromatin Architecture, and Positional Mapping

The cis-eQTL analysis identified several significant SNPs associated with specific tissues, with notably low False Discovery Rates (FDR). Several SNPs in *UQCC1* on chromosome 20 were significantly associated with Cells_Cultured_fibroblasts, with *p* value of 1.25 × 10^-68^, and FDR of 5.39 × 10^-58^. SNPs in *CPNE1* on chromosome 20 were significantly associated with Whole_Blood, which showed *p* values of 1.39 × 10^-50^, and FDR of 7.07 × 10^-188^. Complete data can be found in Supplementary Table 3. Chromatin interaction analysis uncovered significant tissue or cell-specific interactions. Genes related to *GDF5* on chromosome 20 included *EPB41L1*, *CNBD2*, *SCAND1*, *PHF20*, *RMD39*. No significant genes were identified on chromosome 9. On chromosome 15, genes associated with *ALDH1A2* included *AQP9, BNIP2*, *GTF2A2*, *GCNT3*, C15ORF31, and CCNB2. The mapped genes by chromatin interaction on the other chromosomes were provided in Figure 4. The integration of cis-eQTL analysis, chromatin interaction analysis, and positional mapping provided a detailed view of the genomic architecture, highlighting how genetic variants influence gene expression in different tissues.

**Fig. 4.**
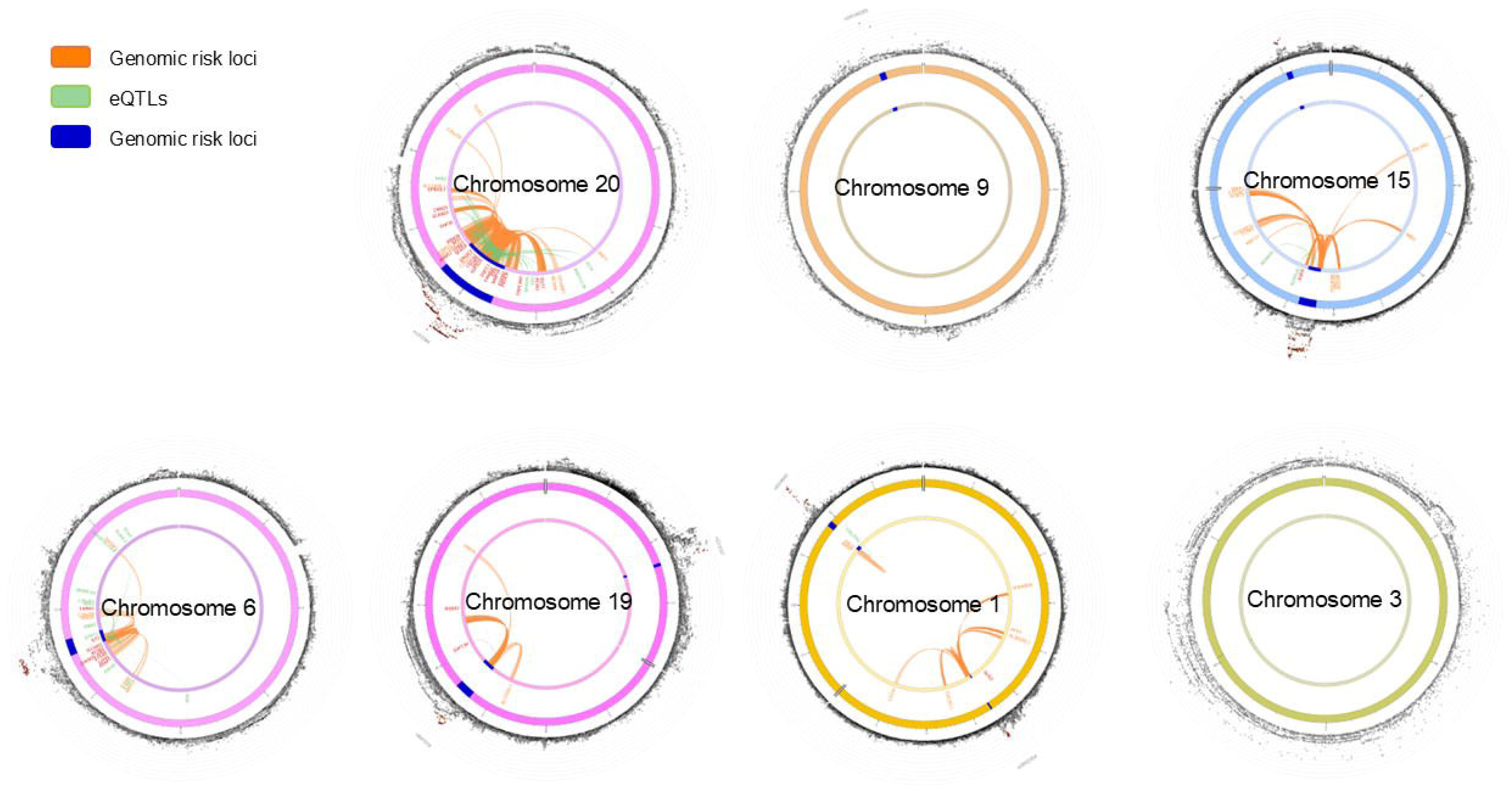
Circos plot illustrating chromatin interactions and eQTL

### Genetic Correlation Analysis using LDSC

In our study on the genetic correlations between knee pain and various traits by Complex-Traits Genetics Virtual Lab, several notable associations emerged. We identified strong genetic correlations between knee pain and several pain-related phenotypes, particularly multisite chronic pain (*r_g_* = 0.72, *p* = 2.06 × 10^-259^), leg pain (*r_g_* = 0.71, *p* = 1.18 × 10^-51^). Furthermore, knee pain exhibited significant positive genetic correlations with various medical conditions and health outcomes, including M15 Polyarthrosis (*r_g_* = 0.86, *p* = 2.57 × 10^-5^), other specific joint derangements (*r_g_* = 0.84, *p* = 1.00 × 10^-9^), and OA (*r_g_* = 0.74, *p* = 9.46 × 10^-57^). These findings are thoroughly detailed in Supplementary Table 4 and visually represented in Figure 5. T Additionally, the genetic correlation for knee pain between male and female participants was estimated (*r_g_* = 0.92, *p* = 3.11 × 10^-39^), potentially offering further insight into the genetic distinctions observed in the sex-stratified secondary GWAS.

**Fig. 5.**
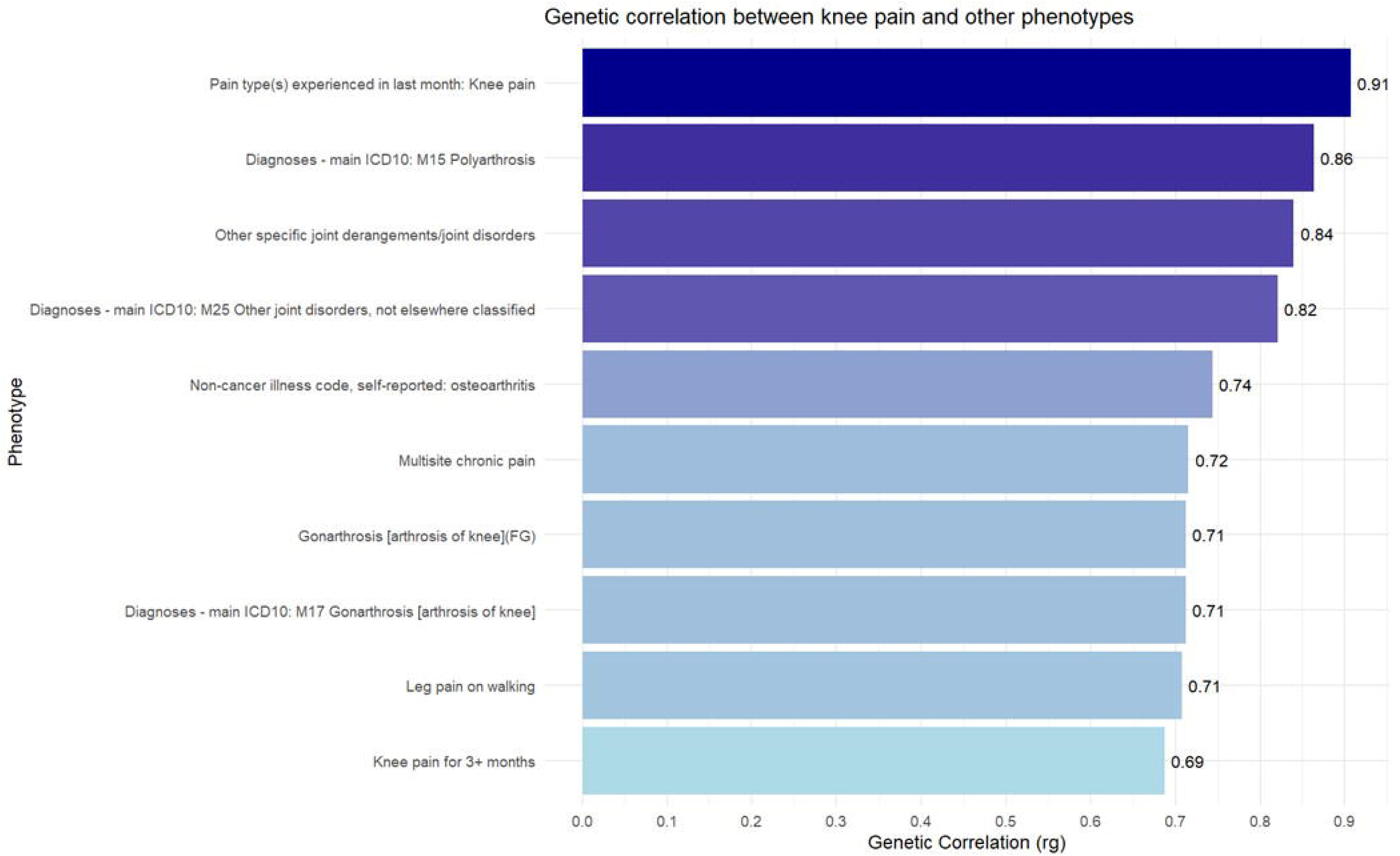
Genetic correlation results for pain intensity using LDSC on Complex-Traits Genetics Virtual Lab. For a full list, see Supplementary Table 4

### TWAS

TWAS conducted across multiple tissue types using GTEx v7 data revealed several statistically significant associations for tissue-specific expression of genes linked to our GWAS-identified SNPs (Supplementary Table 5). After applying the Bonferroni correction for multiple comparisons, the gene *SUPT3H* showed significant expression-trait associations in multiple brain regions, including the nucleus accumbens basal ganglia (Z = 4.40, *p* = 1.08 × 10^-5^), frontal cortex BA17 (Z = 4.16, *p* = 3.21 × 10^-5^), and cerebellum (Z = 3.78, *p* = 1.58 × 10^-4^). Similarly, *RBM6* demonstrated significant associations across various tissues, including spleen (Z = −3.47, *p* = 5.21 × 10^-4^), heart left ventricle (Z = −3.41, *p* = 6.42 × 10^-4^) and thyroid (Z = −3.30, *p* = 9.56 × 10^-4^). The gene *ILF3* showed a significant negative association in nerve tibial (Z = −4.23, *p* = 2.39 × 10^-5^) and esophagus mucosa (Z = −4.06, *p* = 4.91 × 10^-5^). The gene *ALDH1A2* showed a significant positive association in the skin of the lower legs exposed to sunlight (Z = 3.51, *p* = 4.41 × 10^-4^). In contrast, genes such as *COL27A1* and *WDR61* displayed fewer significant associations across the tissues analyzed. *COL27A1* showed marginal significance in the cells transformed fibroblasts (Z = −2.63, *p* = 8.5 × 10^-3^), while *WDR61* had a non-significant association in the adipose visceral omentum (Z = 0.80, *p*= 0.42).

### PheWAS

The GWAS ATLAS platform was leveraged to conduct a PheWAS to explore the phenotypes associated with significantly linked SNPs (rs143384, rs919642, rs56076919, rs34148285, rs3892354, rs687878. rs687878, rs145408100) and their corresponding genes (*GDF5, COL27A1, ALDH1A2, COL11A2, ILF3, HSD11B1, GLIS3, SUPT3H, RBM6*, *WDR61*). The findings pointed to robust associations between rs143384 and rs687878 SNPs with height, exhibiting *p* values of 1 × 10^-300^ and 2.8 × 10^-27^, respectively. Rs919642, rs56076919 and rs3892354 demonstrated strong associations with OA, exhibiting *p* values of 8.55 × 10^-15^, 1.84 × 10^-5^ and 1.78 × 10^-8^. Furthermore, SNP rs3892354 displayed notable associations with thyroid-stimulating hormone (*p* = 9.98 ×10^-12^), male-specific factors - hair/balding pattern (*p* = 2.68 × 10^-10^) and type 2 Diabetes (p = 8.68 × 10^-10^). Regarding gene associations, the *GDF5*, *COL27A1* and *ILF3* genes displayed strong correlations with standing height, showing *p* = 5.28 × 10^-110^, 1.16 × 10^-24^ and 3.00 × 10^-27^, respectively. The *COL11A2* gene showed significant associations with multiple conditions such as rheumatoid arthritis (*p* = 7.74 × 10^-31^), celiac disease (*p* = 2.13 × 10^-24^) and type 1 diabetes (*p* = 1.29 × 10^-21^). A detailed list of traits that passed the Bonferroni correction can be found in Supplementary Table 6. Visual representations of the phenotypes associated with the SNPs and genes are provided in Supplementary Figures 6 and 7.

### MR of Knee Pain and Other Phenotypes

We conducted MR analyses to investigate potential causal relationships between knee pain and various phenotypes, including leg pain on walking, polyarthrosis, other specific joint derangements, and OA. Table 4 provides detailed information on the GWAS datasets used in this study, sourced from the IEU database. We utilize GWAS summary data from IEU, including ‘Leg pain on walking’ with 151,553 participants (33,509 cases and 118,044 controls), ‘Diagnoses - main ICD10: M15 Polyarthrosis’ with 361,194 participants (1,264 cases and 359,930 controls), ‘Other specific joint derangements/joint disorders’ with 361,194 participants (7,943 cases and 353,251 controls), and ‘Non-cancer illness code, self-reported: osteoarthritis’ with 462,933 participants (38,472 cases and 424,461 controls).

**Table 4.**
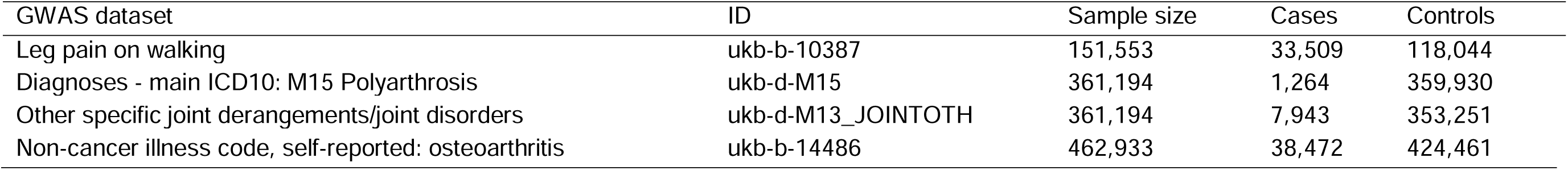
Comprehensive Details of IEU GWAS Datasets for MR Analyses.

In the forward causality analysis, knee pain was examined as the exposure to assess its potential impact on outcomes such as leg pain on walking, polyarthrosis (ICD-10: M15), other specific joint derangements (ICD-10: M13), and osteoarthritis. The Inverse Variance Weighted (IVW) method consistently revealed significant associations (*p* < 0.05), suggesting a strong causal relationship between knee pain and these outcomes. For instance, knee pain was associated with an increased risk of leg pain on walking, with an odds ratio (OR) of 1.39 (95% CI 1.19 to 1.62, *p* = 2.53 ×10^-5^). Similarly, knee pain was found to significantly increase the risk of polyarthrosis (OR = 1.03, 95% CI 1.01 to 1.04, *p* = 2.2 ×10^-4^), other specific joint derangements (OR = 1.06, 95% CI 1.02 to 1.09, *p* = 2.4 ×10^-3^), and osteoarthritis (OR = 1.31, 95% CI 1.15 to 1.49, *p* = 4.68 ×10^-5^). These findings were robust across different MR methods, including the Weighted Median and Simple Mode methods, though the MR Egger method, which accounts for potential pleiotropy, did not always reach statistical significance, indicating the possibility of pleiotropic effects influencing some of the causal estimates.

Conversely, the reverse causality analysis explored whether genetically predicted leg pain on walking, polyarthrosis, other specific joint derangements, and osteoarthritis might causally influence the risk of developing knee pain. The IVW method provided compelling evidence for a significant reverse causal relationship between leg pain on walking and knee pain (OR = 1.28, 95% CI 1.22 to 1.35, *p* = 3.83 ×10^-21^), suggesting that leg pain on walking increases the likelihood of knee pain. Similarly, osteoarthritis was shown to significantly elevate the risk of knee pain (OR = 2.26, 95% CI 1.62 to 3.17, *p* = 1.86 ×10^-6^), further underscoring the bidirectional nature of the relationship between these conditions. However, the reverse analyses for polyarthrosis and other specific joint derangements did not produce statistically significant results, indicating that these conditions may not have a direct causal impact on knee pain. Detailed results and figures are presented in Supplementary Figure 8 and Supplementary Table 7.

## Discussion

In this research of knee pain, utilizing the UK Biobank dataset, we identified ten significant genetic loci, eight of which are novel. The study employed a novel case/control definition based on self-reported knee pain in the past month versus no knee pain in the past month. Notably, seven of the ten significant loci were successfully replicated in the FinnGen cohort. In the sex-stratified GWAS, we observed differences in the prevalence of genetic variants associated with knee pain between males and females, identifying three loci in females and four loci in males.

In the primary GWAS, the locus within the *GDF5* gene on chromosome 20 exhibited the most significant association with knee pain, with the top SNP being rs143384 (*p* = 4.68 × 10^-19^). The second most significant locus located in the *COL27A1* region on chromosome 9, with the top SNP identified as rs919642 (*p* = 1.25 × 10^-17^). The *GDF5* gene, located on chromosome 20, encodes growth differentiation factor 5, a member of the bone morphogenetic protein family, which is essential for the development and repair of bone and cartilage. Variants in *GDF5* have been consistently linked to OA and other joint disorders due to their role in chondrogenesis and joint formation(Pregizer et al. 2018). The SNP rs143384, identified as the top SNP in our study, has been previously reported to influence *GDF5* expression levels, contributing to OA susceptibility by affecting cartilage homeostasis (Mikic 2004; Pregizer et al. 2018). Similarly, the *COL27A1* gene, which encodes collagen type XXVII alpha 1 chain, is crucial for cartilage structure and function. Collagen type XXVII is a minor collagen component that plays a significant role in the early stages of cartilage development and in maintaining the integrity of the extracellular matrix (Gonzaga-Jauregui et al. 2020). Variants in the *COL27A1* region, such as the SNP rs919642 identified in our study, may affect collagen fibril formation and stability, thereby influencing the structural integrity of cartilage and contributing to knee pain and OA (Gonzaga-Jauregui et al. 2015). Previous GWAS research of knee pain has also identified these two loci, further supporting our findings and their relevance to knee pain (Meng et al. 2019).

The most significant novel locus identified in our study is within the *ALDH1A2* gene area on chromosome 15, with the top SNP being rs34291892 (*p* = 1.69 × 10^-9^). *ALDH1A2* encodes aldehyde dehydrogenase 1 family member A2, an enzyme critical for the biosynthesis of retinoic acid, a metabolite of vitamin A that regulates gene expression during cell differentiation, proliferation, and apoptosis. Retinoic acid is essential for maintaining cartilage homeostasis and joint integrity (Napoli 2012). Variants in *ALDH1A2* have been associated with various skeletal and cartilage-related disorders, indicating that changes in retinoic acid synthesis can impact cartilage health. Retinoic acid influences the expression of matrix metalloproteinases (MMPs), enzymes that degrade extracellular matrix components and contribute to cartilage breakdown in OA (Mehana et al. 2019; Pulik et al. 2023). Dysregulation of MMP activity due to altered retinoic acid levels can lead to cartilage degradation and subsequent knee pain. *ALDH1A2* expression is also linked to inflammatory pathways involved in joint pain and degeneration. Studies have shown that retinoic acid modulates inflammatory cytokine production, affecting the inflammatory environment of the joint and exacerbating pain symptoms (Davies et al. 2009). The SNP identified in our study may influence *ALDH1A2* expression or activity, thereby impacting these pathways and contributing to the development of knee pain. Additionally, *ALDH1A2* plays a crucial role in chondrocyte differentiation and maintenance, highlighting its importance in cartilage biology. Disruptions in *ALDH1A2* function could impair chondrocyte differentiation, leading to compromised cartilage integrity and increased susceptibility to knee pain and OA. Furthermore, previous GWAS have reported associations between this gene and knee OA, supporting a potential mechanistic link between *ALDH1A2* and the pathophysiology of knee pain (Tachmazidou et al. 2019; Henkel et al. 2023).

In our GWAS, we identified another significant novel association between knee pain and the *ILF3* gene on chromosome 19, with the top SNP rs56764248 (*p* = 1.29 × 10^-14^). *ILF3*, also known as Interleukin Enhancer Binding Factor 3, is a protein-coding gene involved in various cellular processes, including mRNA stabilization and regulation of gene expression. This gene has been previously reported to be associated with rheumatoid arthritis by several GWAS studies (Laufer et al. 2019; Ha et al. 2021; Saevarsdottir et al. 2022). ILF3’s role in rheumatoid arthritis suggests it may contribute to the inflammatory pathways involved in knee pain. Additionally, *ILF3* is involved in the NF-κB signaling pathway, a critical regulator of inflammation and immune response. Dysregulation of this pathway can lead to chronic inflammation and joint damage, further linking *ILF3* to knee pain (Nazitto et al. 2021). *ILF3* could play a role in the immune response and inflammation, supporting its potential impact on joint health and pain (Nazitto et al. 2021). The identification of *ILF3* as a gene associated with knee pain in our GWAS underscores its potential role in the inflammatory processes that contribute to joint pain and degeneration. Understanding the mechanisms by which *ILF3* influences inflammation and cartilage degradation could provide new insights into therapeutic targets for knee pain and related conditions.

We identified four additional significant novel associations with knee pain: *COL11A2* on chromosome 6 with the top SNP rs56076919 (*p* = 9.82×10^-12^), *GLIS3* on chromosome 9 with the top SNP rs3892354 (*p* = 1.29 × 10^-8^), *SUPT3H* on chromosome 6 with the top SNP rs687878 (*p* = 2.38 × 10^-8^), and *RBM6* on chromosome 3 with the top SNP rs687878 (*p* = 3.17 × 10^-8^). These genes have previously been associated with knee OA in other GWAS studies, indicating their potential roles in OA pathogenesis (Tachmazidou et al. 2019; Hollis et al. 2023). *COL11A2* encodes the a2(Xl) component of collagen Xl, is linked to autosomal human osteochondrodysplasia(Vikkula et al. 1995). *GLIS3* encodes a protein involved in transcriptional regulation and cellular homeostasis. It has been linked to OA susceptibility, potentially due to its role in chondrocyte differentiation and cartilage integrity (Casalone et al. 2018). *SUPT3H* is involved in transcriptional regulation and chromatin remodeling, and its dysregulation can lead to altered gene expression profiles in joint tissues, contributing to cartilage degradation and inflammation (Rice et al. 2018). Additionally, *SUPT3H* has been reported associated with back pain, suggesting a shared mechanism between different pain types (Bjornsdottir et al. 2022). *RBM6* encodes an RNA-binding protein that plays a role in mRNA processing and stability. Its association with OA suggests it may influence the expression of genes involved in cartilage homeostasis and response to mechanical stress, contributing to knee pain (Castaño-Betancourt et al. 2016). The identification of these genes underscores their potential involvement in the molecular mechanisms underlying knee pain and OA, offering new insights into the pathogenesis for these conditions.

Among the additional loci identified in our primary GWAS, the genes *HSD11B1*, and *WDR61* exhibited significant associations. These genes had not been previously linked to knee disorders in genetic studies. However, the presence of multiple significant SNPs in these loci suggests their potential importance. These regions are gene-rich, indicating complex regulatory mechanisms. Specifically, the *HSD11B1* gene has been implicated in cartilage metabolism in prior research, positioning it as a potential key player in the onset or progression of knee disorders (Kragl et al. 2022). Despite these findings, further research is required to validate these associations and elucidate their precise roles in the knee pain phenotype.

In our sex-stratified GWAS of knee pain, we identified distinct genetic loci associated with knee pain in males and females, indicating potential sex-specific genetic mechanisms underlying this condition. For males, significant loci included *ALDH1A2* on chromosome 15 (rs2899611, *p* = 2.77 × 10^-11^), *COL27A1* on chromosome 9 (rs891720, *p* = 5.55 × 10^-11^), *ILF3* on chromosome 19 (rs2742313, *p* = 4.19 × 10^-9^), and *MBNL1* and *RP11-362A9.3* on chromosome 3 (rs2019689, *p* = 6.51 × 10^-9^). For females, significant loci included *GDF5* on chromosome 20 (rs143384, *p* = 1.70 × 10^-15^), *COL11A2* on chromosome 6 (rs56076919, *p* = 9.82 × 10^-12^), and *COL27A1* on chromosome 9 (rs919642, *p* = 1.45 × 10^-8^). A notable observation is the shared association with the *COL27A1* gene in both males and females, although the top SNPs differ. This suggests that *COL27A1* may play a crucial role in knee pain pathogenesis across sexes, potentially affecting cartilage structure and function. In males, the involvement of the *ALDH1A2* and *ILF3* genes in the primary GWAS results suggests that these genes are primarily contributing to knee pain in males, as they did not appear significantly in the female-specific GWAS. Additionally, the male-specific GWAS indicated that *MBNL1* and the non-coding RNA *RP11-362A9.3* on chromosome 3, which are involved in RNA splicing and regulation, may indicate a complex post-transcriptional regulatory mechanism in male knee pain (Ho et al. 2004). In females, the *GDF5* gene showed a highly significant association, highlighting its importance in female knee pain. The association with *COL11A2*, another collagen gene, suggests that extracellular matrix composition and integrity are critical factors in female knee pain (Shi et al. 2010). These findings underscore both shared and distinct genetic mechanisms underlying knee pain in males and females. While the shared involvement of *COL27A1* points to common pathways in cartilage structure, the unique associations with *ALDH1A2*, *ILF3*, and *MBNL1* in males, and *GDF5* and *COL11A2* in females, suggest sex-specific pathways influencing knee pain. The calculated genetic correlation between males and females in knee pain (*r_g_* = 0.92) may further support this concept. Further research is needed to elucidate the precise roles of these genes and their interactions in knee pain pathophysiology, potentially leading to sex-specific therapeutic strategies.

In our study, we employed a novel case/control definition, comparing individuals with knee pain to those with no knee pain within the last month, which may capture more relevant mechanisms underlying knee pain. Compared to previous GWAS on knee pain using the UK Biobank cohort, this study discovered eight new loci in the primary GWAS (Meng et al. 2019). In the tissue expression analysis, knee pain showed a relatively high correlation with brain tissues. This suggests that the genetic determinants of knee pain may influence neurological pathways involved in pain perception and processing. Previous studies have identified significant gene expression correlations between brain tissues and chronic pain conditions, underscoring the role of the nervous system in pain modulation (Crofford 2015; Araki and Mimura 2017). This aligns with findings that genes associated with pain often exhibit co-expression in brain tissues, highlighting the importance of central nervous system pathways in the manifestation of pain symptoms (Yam et al. 2018).

Through TWAS, the significant associations observed for certain genes across multiple tissues suggest that these genes may play critical roles in the biological processes contributing to knee pain. *SUPT3H* emerged as one of the most consistently associated genes, showing significant expression across various brain regions, including the nucleus accumbens basal ganglia, frontal cortex BA17 and cerebellum. These findings are particularly noteworthy given the central role of the brain in pain perception and modulation. The consistent expression of *SUPT3H* in these regions may indicate its involvement in the central nervous system’s pathways that process pain signals, potentially contributing to the chronicity or severity of knee pain(Ren and Dubner 1999). This gene’s expression in the pituitary gland further suggests a possible interaction with the endocrine system, which could influence pain through neuroendocrine pathways. *RBM6* also showed significant associations across a wide range of tissues, including both glands and brain tissues. The gene’s expression in thyroid and pituitary suggests that *RBM6* may influence knee pain through mechanisms that involve both endocrine and neural pathways. The involvement of *RBM6* in these tissues points to a possible role in systemic processes such as hormone regulation or neuroendocrine signaling, which could contribute to the perception or modulation of pain. The consistent associations observed for *SUPT3H* and *RBM6* across various tissues suggest that these genes could be contributors to the development or persistence of knee pain, potentially through mechanisms involving both the central nervous and endocrine system.

The genetic correlations between knee pain and several other phenotypes suggest that some joint conditions share common genetic determinants. The three phenotypes most highly correlated with knee pain were polyarthritis (*r_g_* = 0.86), joint disorders (*r_g_* = 0.84), and OA (*r_g_* = 0.77), indicating a strong relationship between knee pain and OA. This supports the use of a knee arthrosis cohort (FinnGen) for our replication. However, since the number of knee pain cases greatly exceeds the number of self-reported knee OA cases (N = 22,204), it is meaningful to treat knee pain as an independent disorder for research purposes, rather than solely as a symptom of knee OA [11]. Our PheWAS of knee pain revealed that the top SNPs and their corresponding genes are significantly associated with various skeletal traits, including hip or knee OA, height, standing height, and sitting height. These findings underscore the interconnected genetic determinants influencing both knee pain and broader skeletal characteristics. The strong associations with hip and knee OA reinforce the shared genetic underpinnings between knee pain and OA, suggesting overlapping biological pathways. Additionally, the links to standing and sitting height highlight the role of these genetic variants in skeletal growth and development, which may affect joint health. These insights emphasize the need to consider pleiotropic effects in genetic studies and suggest that the genetic architecture of knee pain involves broader skeletal morphology.

Furthermore, the MR analyses demonstrated strong evidence for a causal relationship between knee pain and leg pain on walking. Our forward causality analysis indicated that genetic predisposition to knee pain significantly increases the risk of leg pain on walking, as evidenced by the Inverse Variance Weighted (IVW) method (OR = 1.39, 95% CI 1.19 to 1.62, *p* = 2.53 × 10^-5^). Interestingly, the reverse causality analysis also revealed that leg pain on walking causally influences the risk of knee pain (OR = 1.28, 95% CI 1.22 to 1.35, *p* = 3.83 × 10^-21^).

This bidirectional relationship suggests that these conditions may share common genetic pathways, and that interventions targeting one condition could potentially mitigate the risk of the other. The mutual influence between knee pain and leg pain on walking highlights the need for an integrated approach in the management of musculoskeletal pain, where addressing knee pain may alleviate leg pain and vice versa. Our forward MR analysis also identified a significant causal link between knee pain and polyarthrosis (OR = 1.03, 95% CI 1.01 to 1.04, *p* = 2.2 × 10^-4^). This finding underscores the role of knee pain as a contributing factor to the development of polyarthrosis, a condition characterized by the degeneration of multiple joints. However, the reverse analysis did not provide significant evidence for polyarthrosis as a direct cause of knee pain, suggesting that while knee pain may predispose individuals to the broader joint degeneration seen in polyarthrosis, the reverse pathway might not be as strong or direct. The analysis also revealed a significant causal association between knee pain and other specific joint derangements (OR = 1.06, 95% CI 1.02 to 1.09, *p* = 2.4 × 10^-3^). This result indicates that knee pain may predispose individuals to a broader spectrum of joint disorders, beyond the knee itself. The lack of significant reverse causality in this context further supports the notion that knee pain may be an upstream factor in the pathogenesis of other joint conditions. One unsurprsing findings of this study is the bidirectional causal relationship between knee pain and osteoarthritis. The forward analysis showed that knee pain significantly increases the risk of developing osteoarthritis (OR = 1.31, 95% CI 1.15 to 1.49, p = 4.68 × 10^-5^), while the reverse analysis demonstrated that osteoarthritis robustly elevates the likelihood of knee pain (OR = 2.26, 95% CI 1.62 to 3.17, *p* = 1.86 × 10^-6^). This bidirectional relationship underscores the intertwined nature of knee pain and osteoarthritis, suggesting that they may reinforce each other in a vicious cycle. Clinically, this highlights the importance of early identification and treatment of both conditions to potentially break this cycle and improve patient outcomes. The shared genetic factors and pathways contributing to both knee pain and osteoarthritis warrant further investigation to develop targeted interventions that can simultaneously address both conditions. These MR findings have important implications for our understanding of the genetic and causal relationships between knee pain and related musculoskeletal conditions.

Although we obtained some positive findings, there are several limitations in our study. In the replication phase, due to the absence of datasets on knee pain, we utilized knee OA as a proxy phenotype since knee pain is a prominent indicator of knee OA. This substitution may introduce some deviations in the replication stage. Additionally, our use of a broader control group, which could include individuals with pain at other body sites, might impact the GWAS results. It’s crucial to recognize that the classification of cases and controls within the UK Biobank is derived from self-reported responses to a specific question targeting knee pain in a questionnaire. This question aimed to assess the presence of knee pain that significantly impacted daily activities within the past month. However, it did not capture detailed information regarding the intensity, frequency, or precise anatomical location of the pain within the knee. As a result, the phenotype created from these responses should be considered as broadly defined. Additionally, the dependence on self-reported data introduces potential biases, such as inaccuracies in recall and personal interpretation of pain experiences. Future research should build upon the insights from the UK Biobank’s pain rephenotyping survey conducted in 2019, which utilized a more detailed questionnaire, to enhance our comprehensive understanding of the topic. Incorporating objective measures of pain and more precise phenotyping could significantly improve the reliability and validity of future GWAS studies on knee pain (https://biobank.ndph.ox.ac.uk/showcase/ukb/docs/pain_questionnaire.pdf).

## Conclusion

In summary, our primary GWAS identified ten genetic loci associated with knee pain, including eight novel ones, using a novel phenotype definition. Our secondary GWAS revealed both shared and distinct genetic variants of knee pain between males and females. These discoveries provide valuable insights into the genetic underpinnings of knee pain.

## Data Availability

The summary statistics from the UK Biobank regarding knee pain will be made available upon publication. Additional data relevant to this study, not included in the article or supplementary materials, can be obtained from the authors upon reasonable request.

## Acknowledgement

This study fully complies with the ethical standards and data protection regulations of the UK Biobank. The research was conducted using data from the UK Biobank under Application Number 89386. The summary statistics from the UK Biobank regarding knee pain will be made available upon publication. Additional data relevant to this study, not included in the article or supplementary materials, can be obtained from the authors upon reasonable request.

## Authors’ Contributions

All authors were involved in developing the analysis plan. YT and QP carried out the GWAS analysis using the UK Biobank data and prepared the initial manuscript. TC and LY were responsible for data preparation. MH and TD contributed critical feedback on the manuscript. WM coordinated the project and provided further feedback.

## Corresponding authors

Correspondence should be directed to Weihua Meng.

## Consent to Publish

All authors have agreed to the publication of this work.

## Ethical Approval

This research received approval from the Ethics Committee of the University of Nottingham Ningbo China.

