## Supplementary figures and images for "A Genome-wide Association Study Identifies Novel Genetic Variants Associated with Knee Pain in the UK Biobank (N = 441,757)"

### Supplementary Figures 1

#### Slide 1
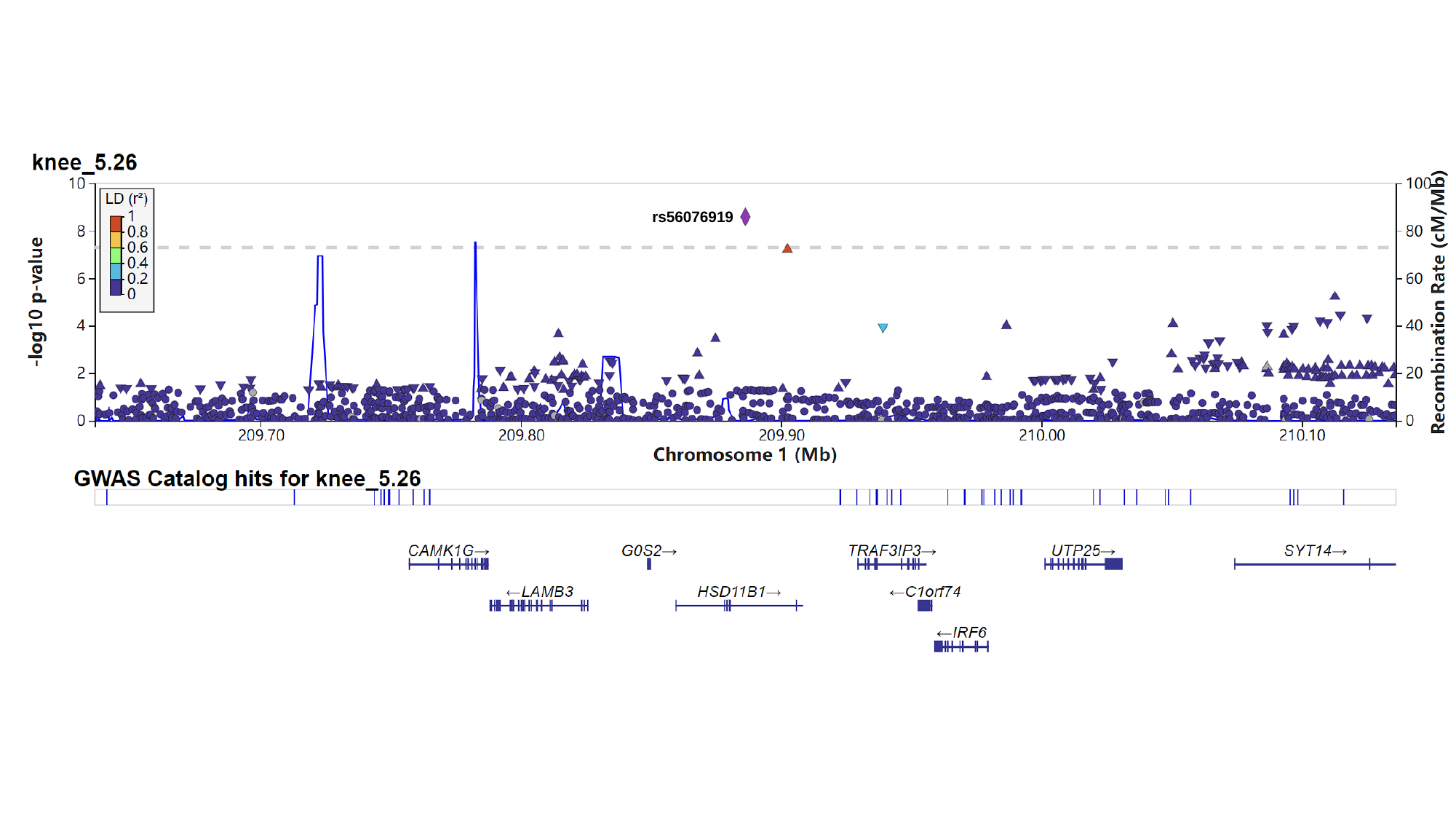

rs56076919

#### Slide 2
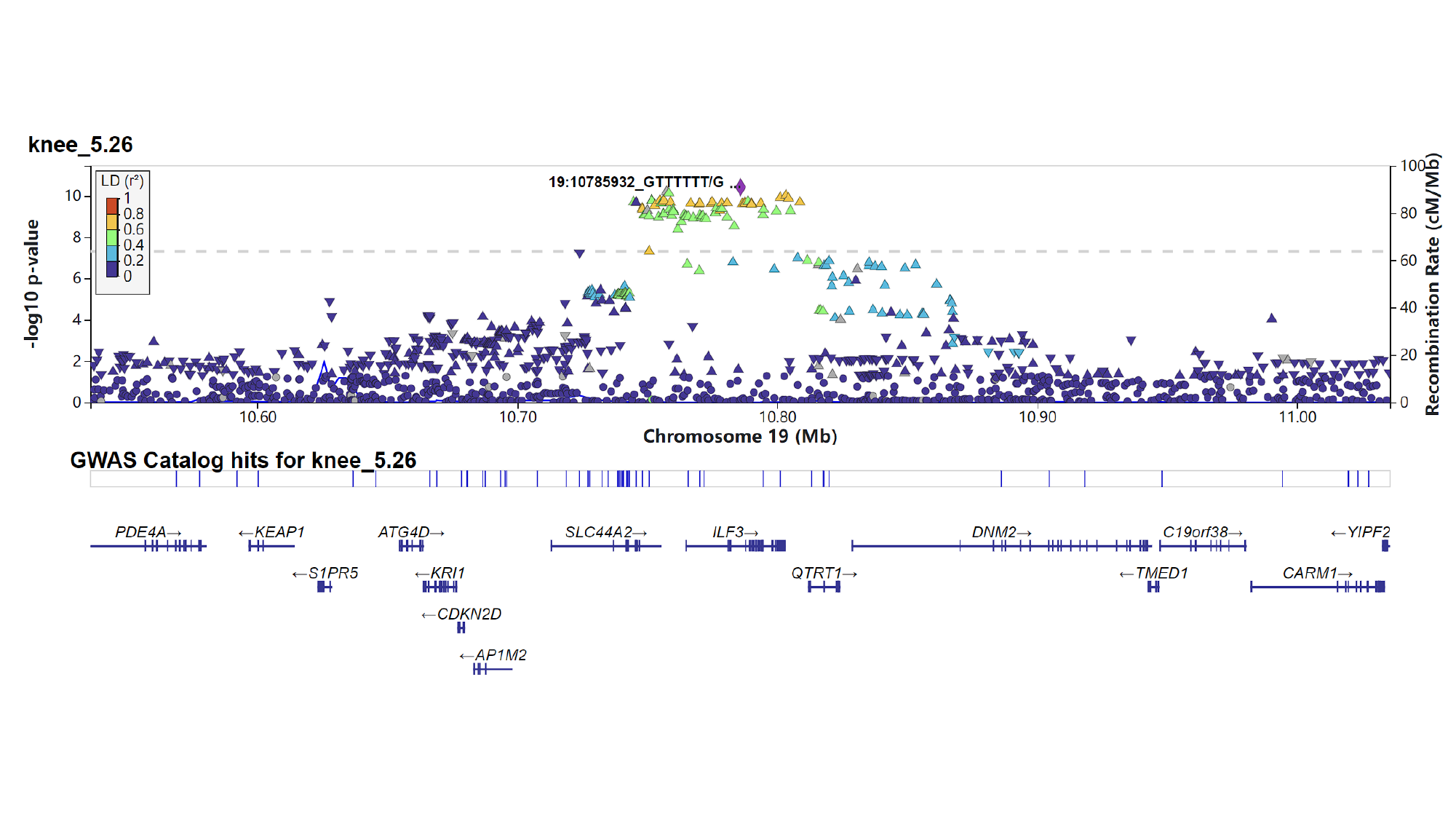

#### Slide 3
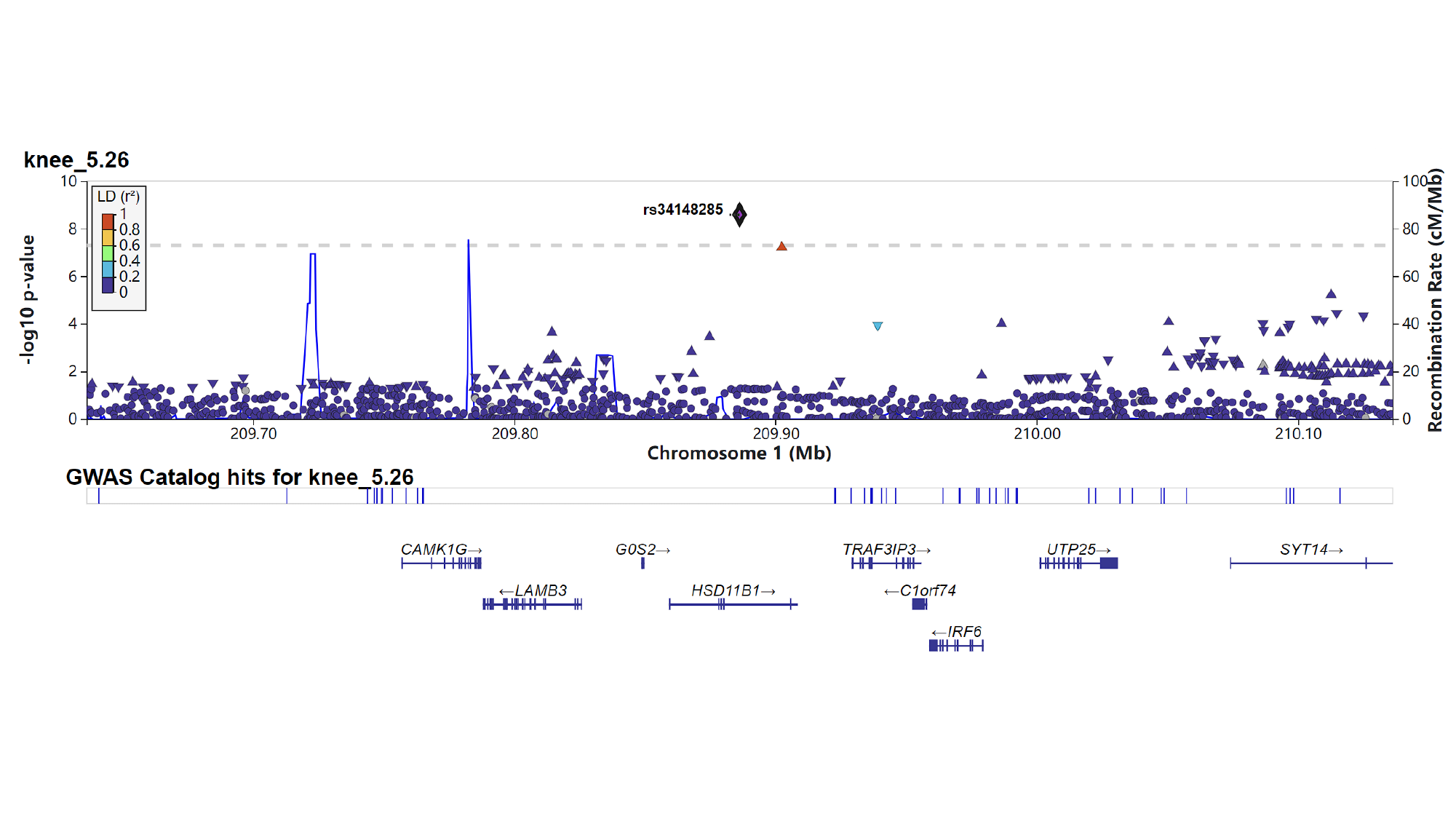

#### Slide 4
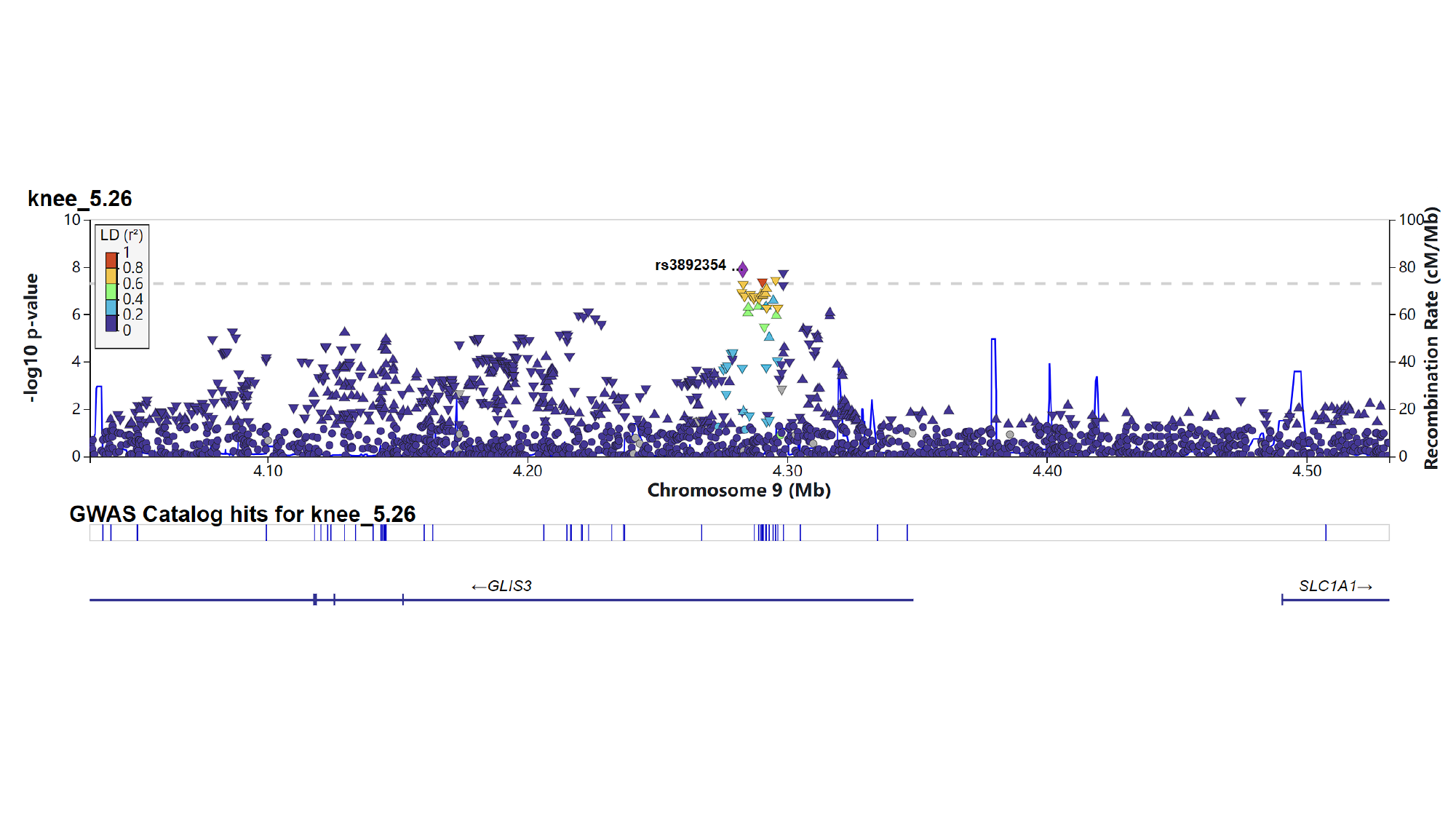

#### Slide 5
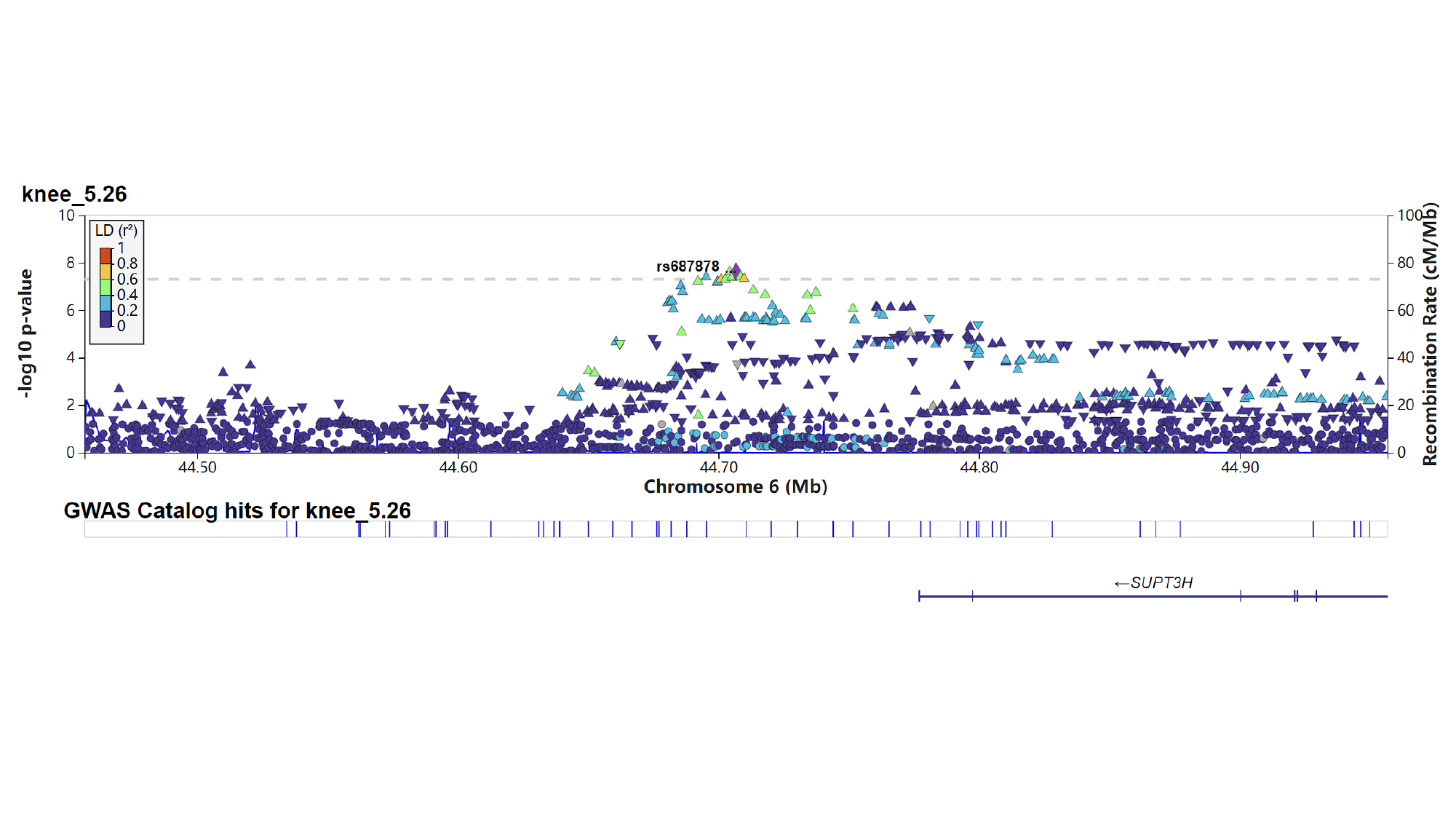

#### Slide 6
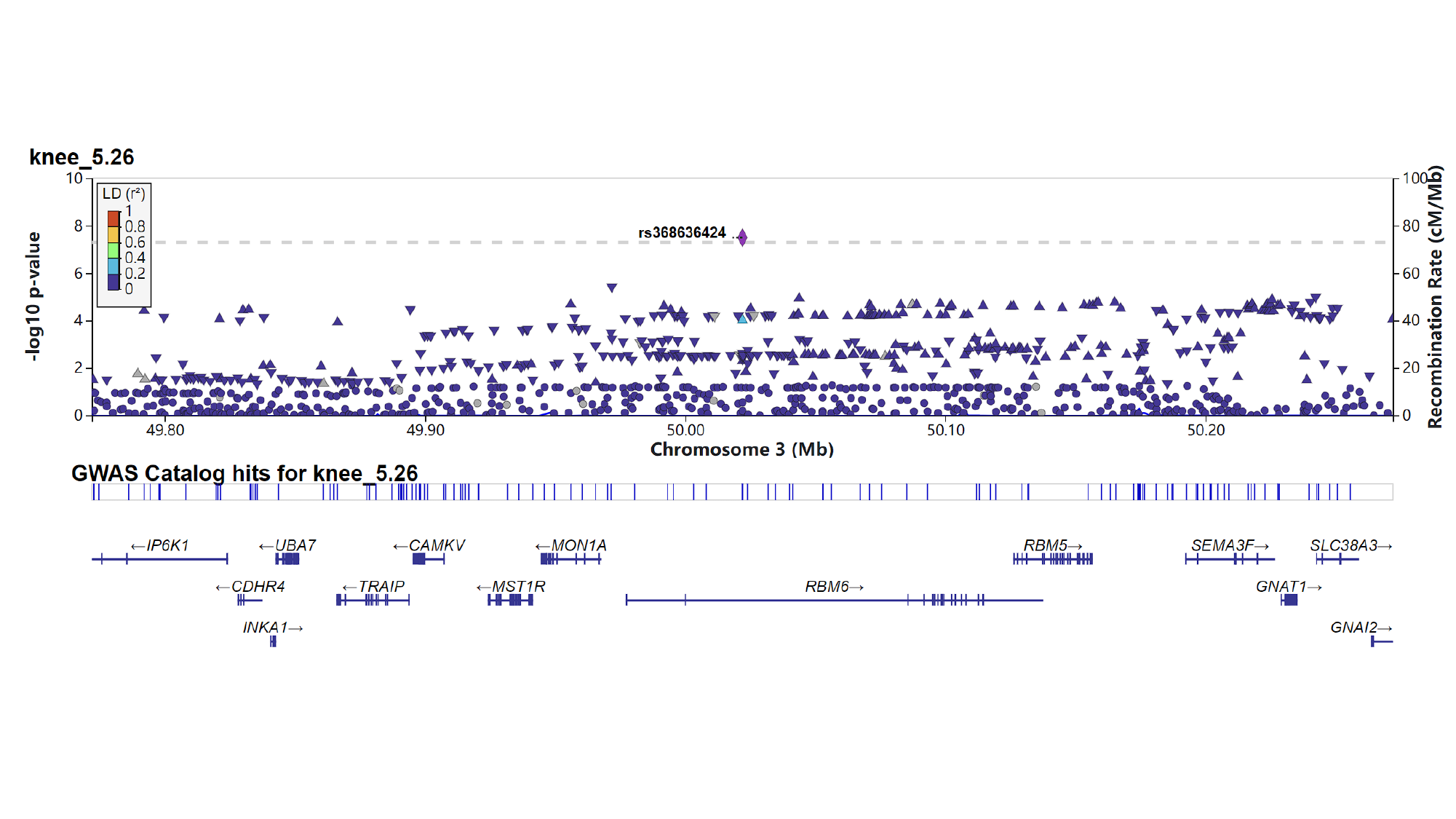

#### Slide 7
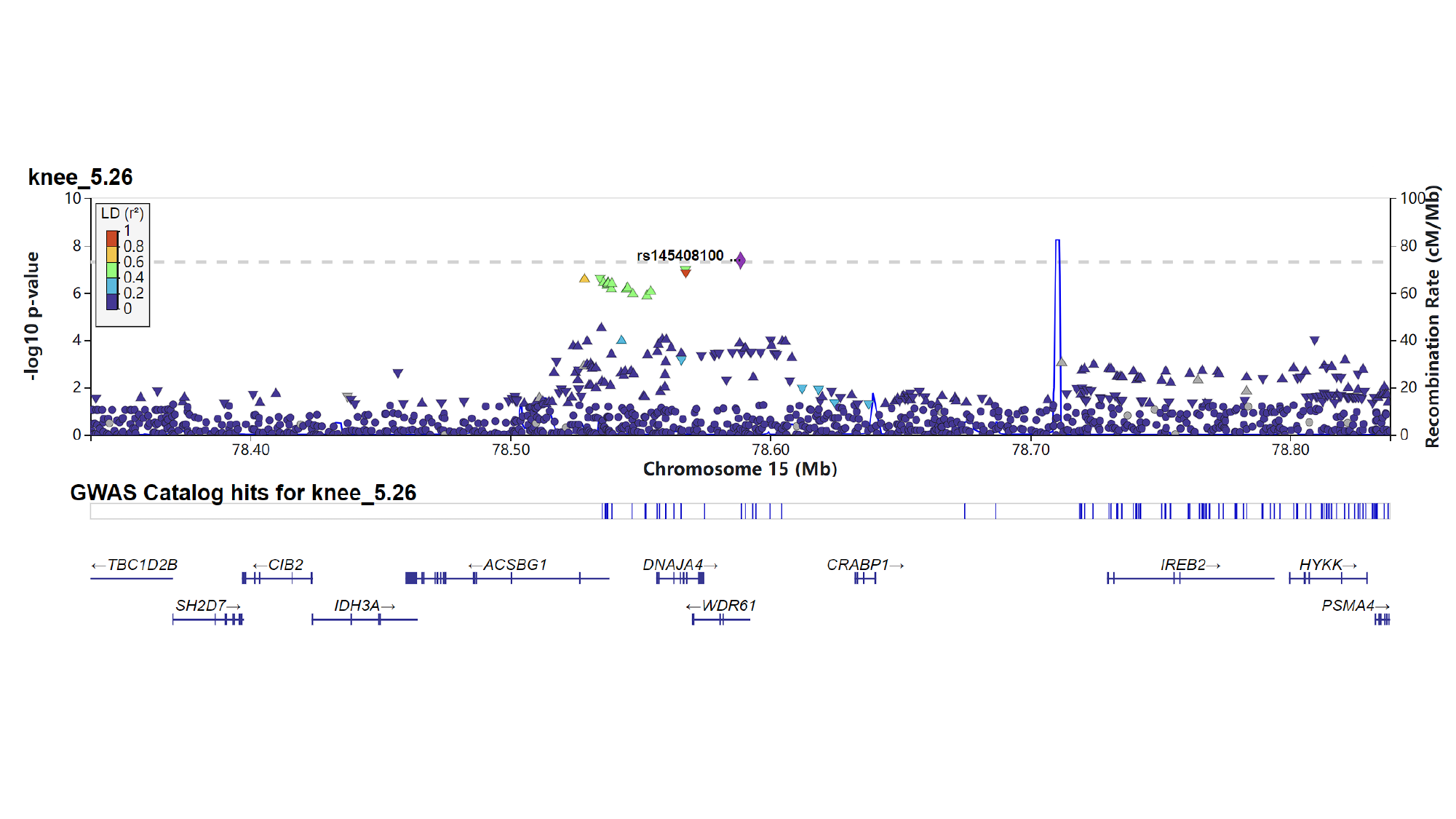

### Supplementary Figures 2

#### Slide 1
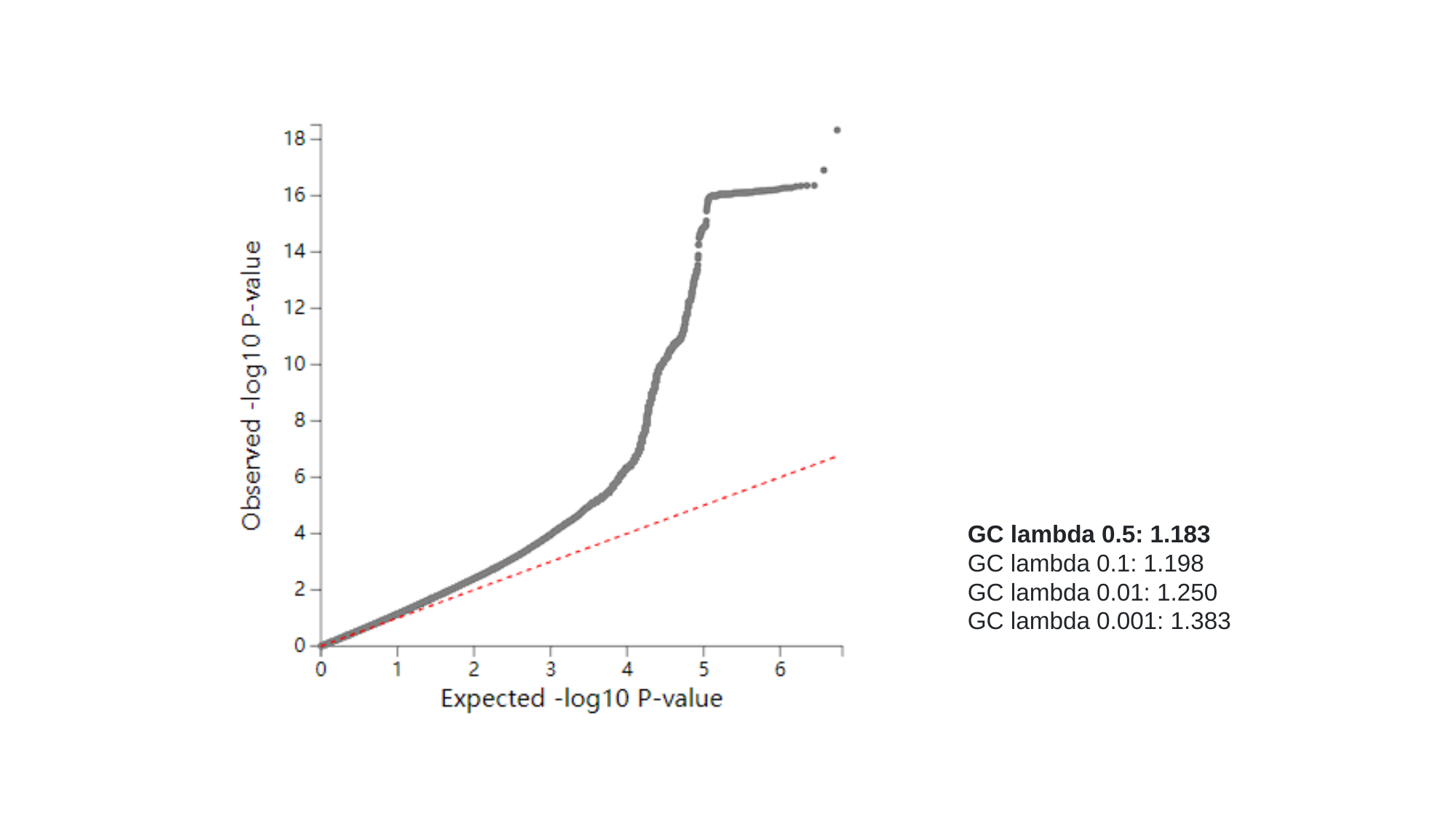

GC lambda 0.5: 1.183
GC lambda 0.1: 1.198
GC lambda 0.01: 1.250
GC lambda 0.001: 1.383

### Supplementary Figures 3

#### Slide 1
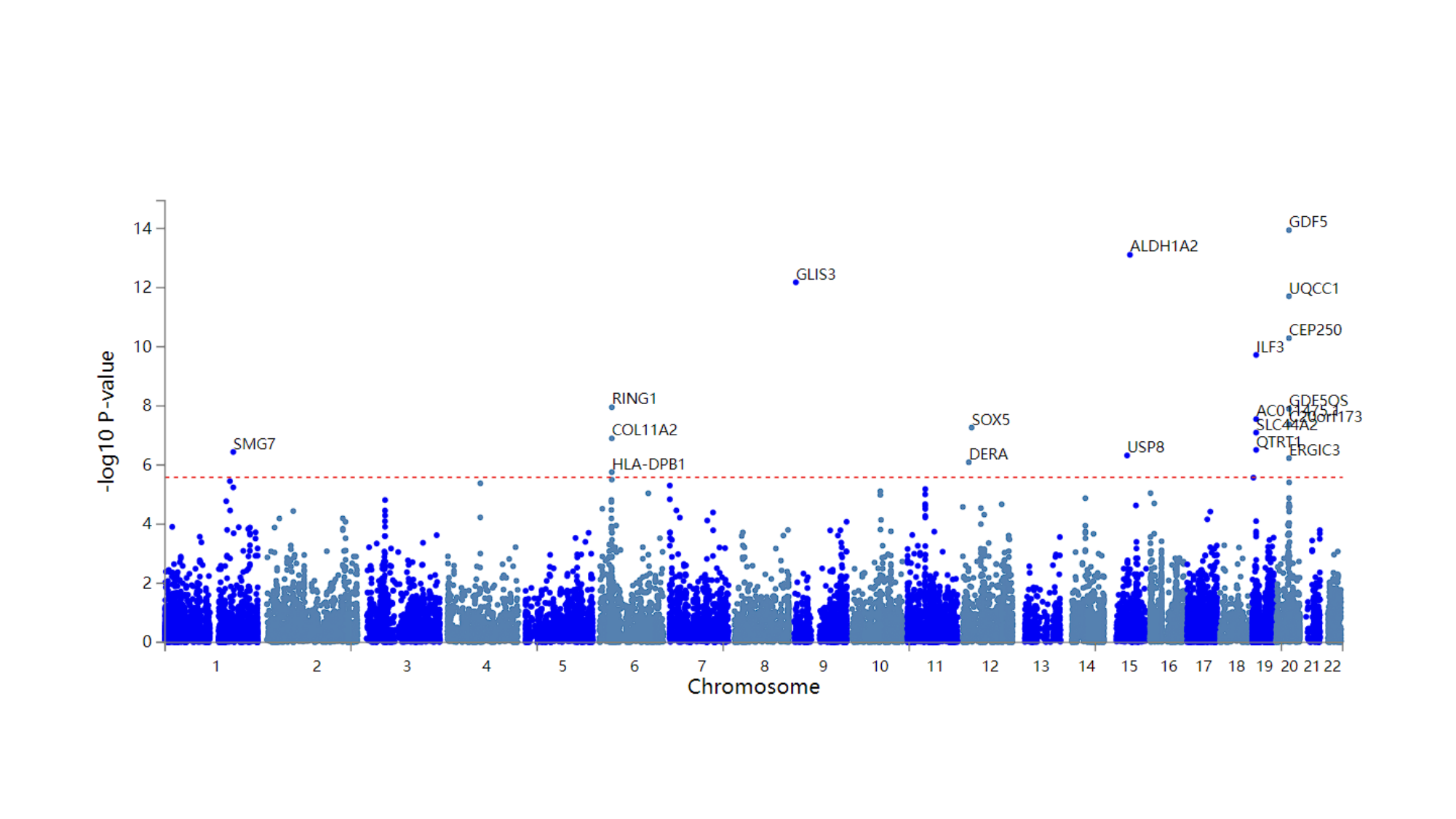

### Supplementary Figures 4

#### Slide 1
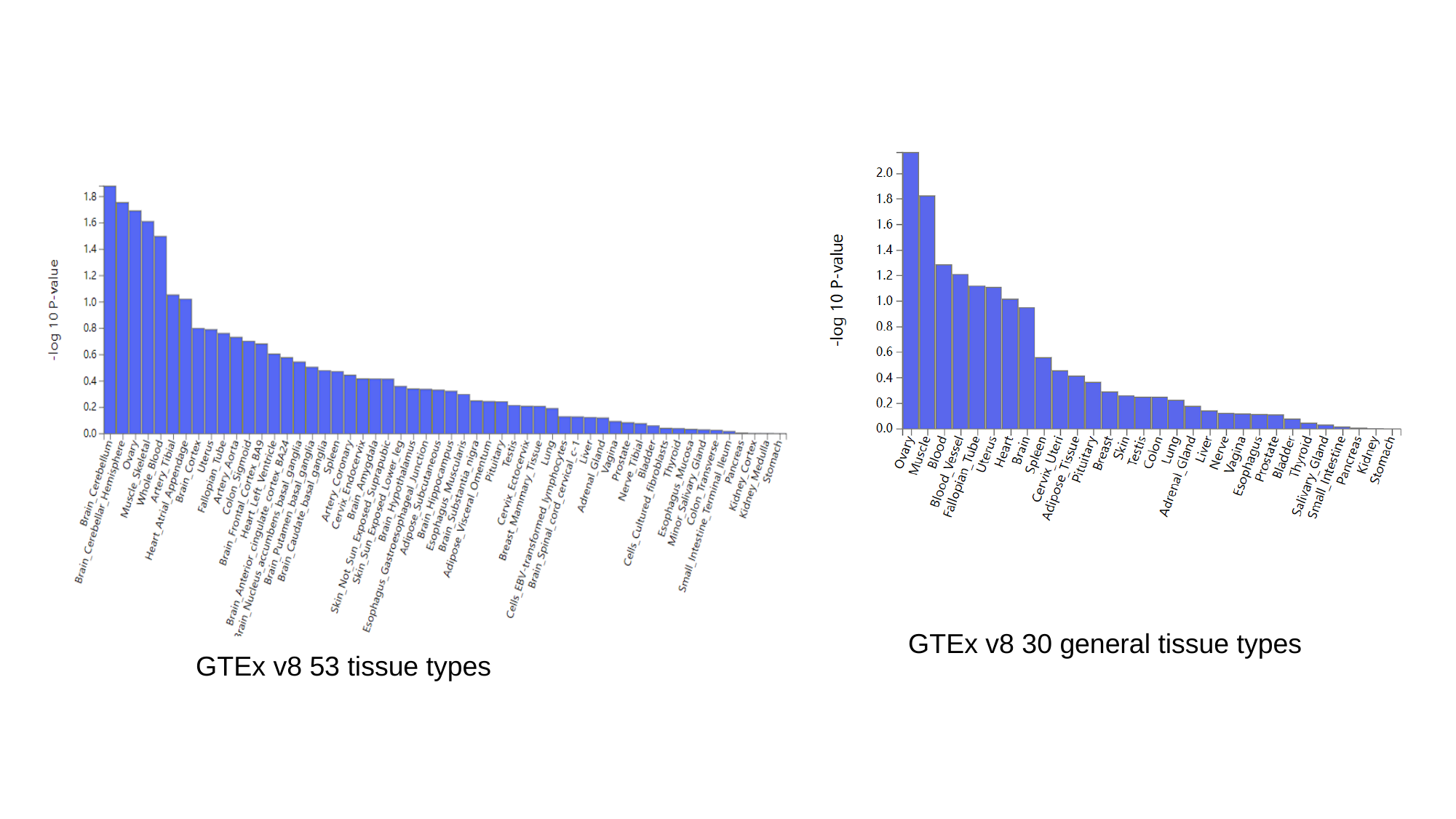

GTEx v8 30 general tissue types
GTEx v8 53 tissue types
