## Supplementary Figures 5 for "A Genome-wide Association Study Identifies Novel Genetic Variants Associated with Knee Pain in the UK Biobank (N = 441,757)"

### Slide 1
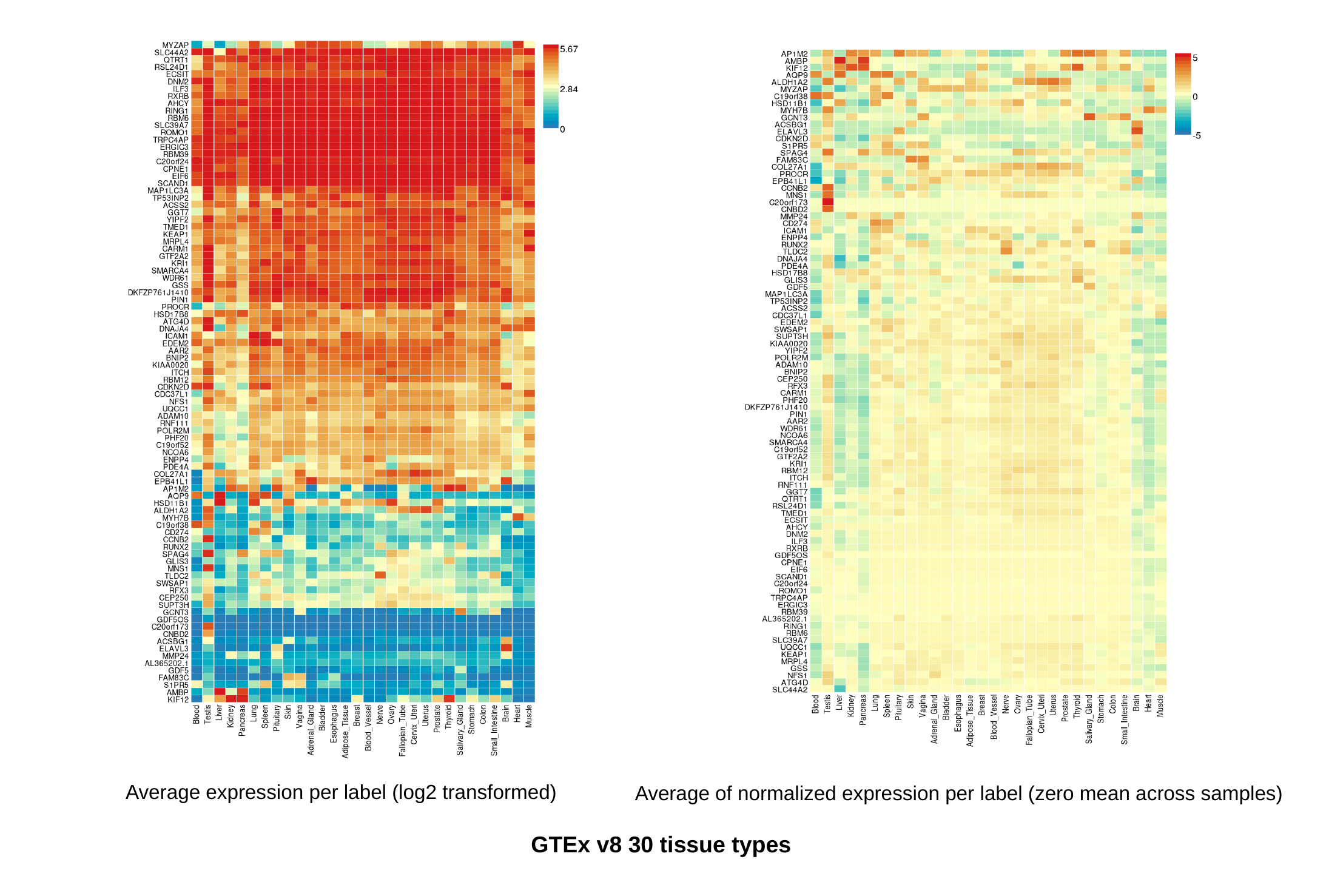

Average expression per label (log2 transformed)
Average of normalized expression per label (zero mean across samples)
GTEx v8 30 tissue types

### Slide 2
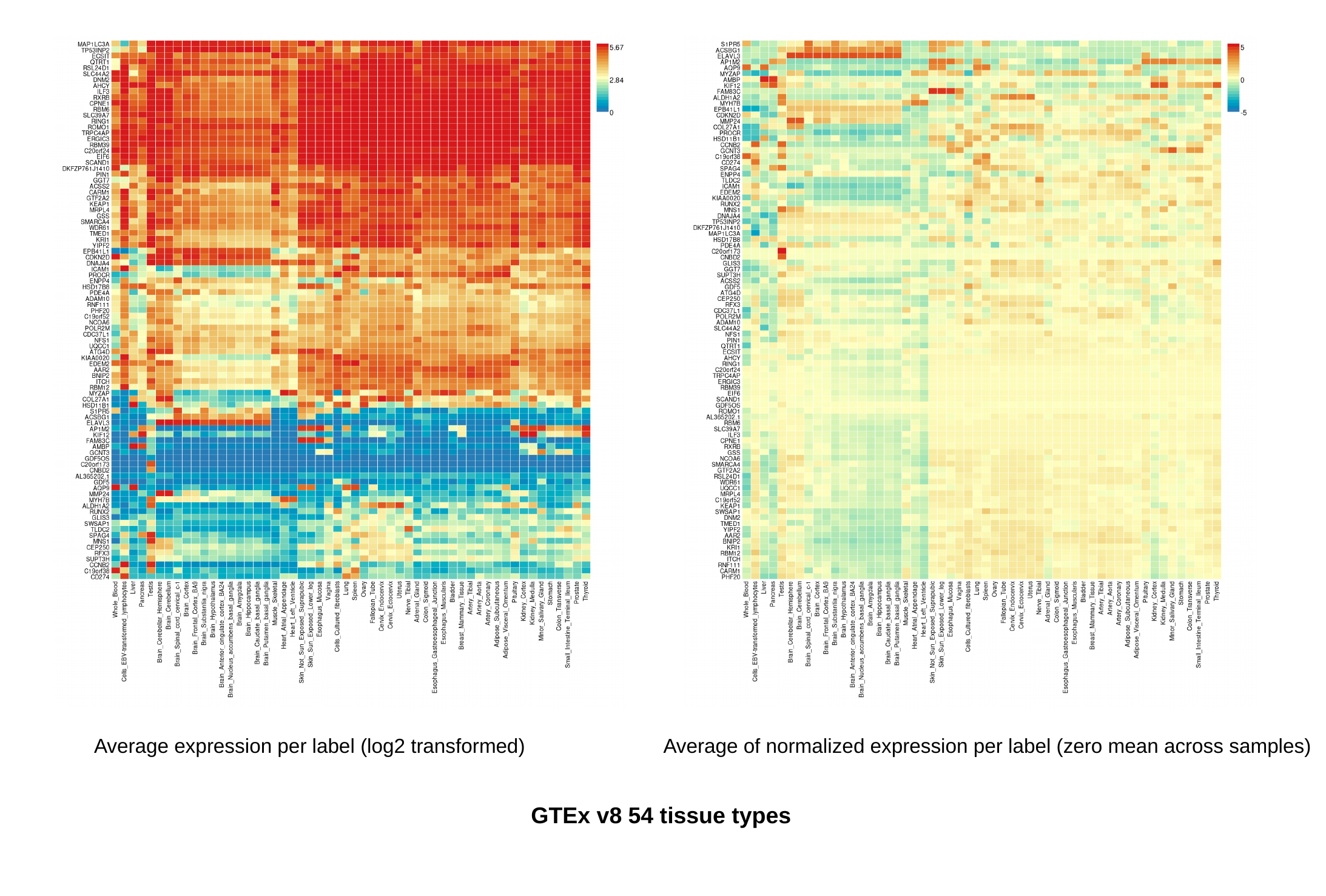

Average expression per label (log2 transformed)
Average of normalized expression per label (zero mean across samples)
GTEx v8 54 tissue types
