## Supplementary Figures 6 for "A Genome-wide Association Study Identifies Novel Genetic Variants Associated with Knee Pain in the UK Biobank (N = 441,757)"

### Slide 1
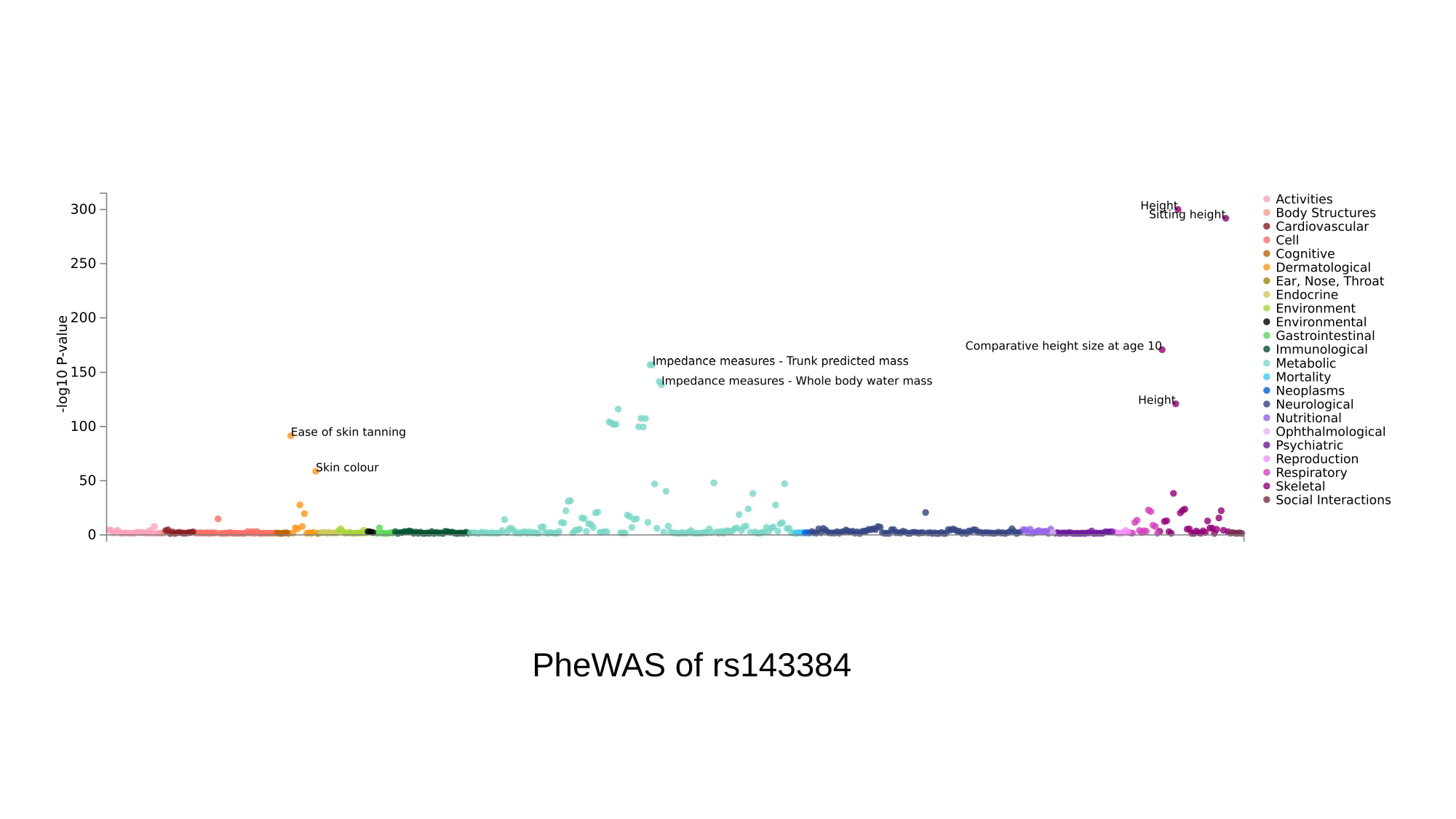

PheWAS of rs143384

### Slide 2
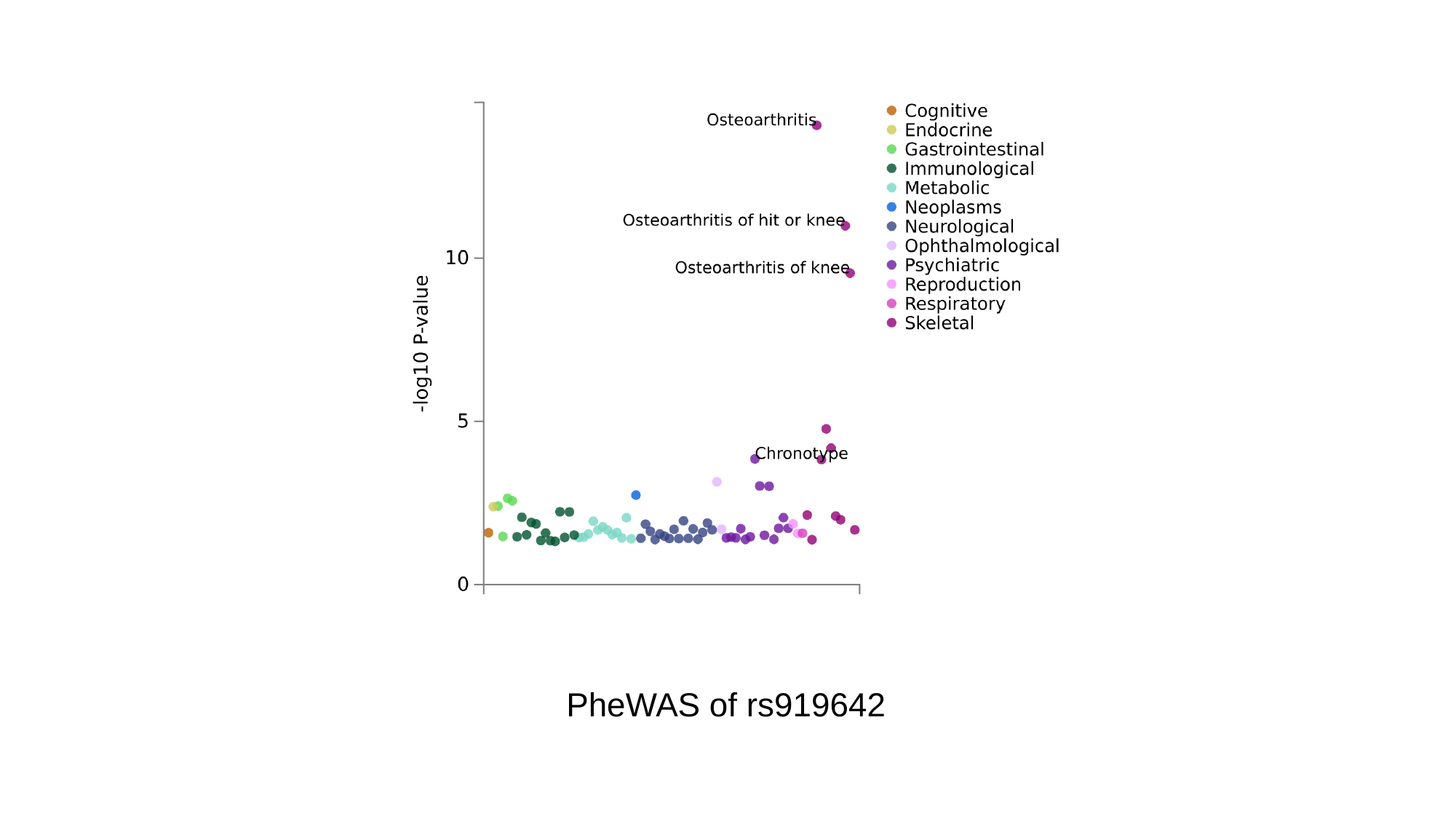

PheWAS of rs919642

### Slide 3
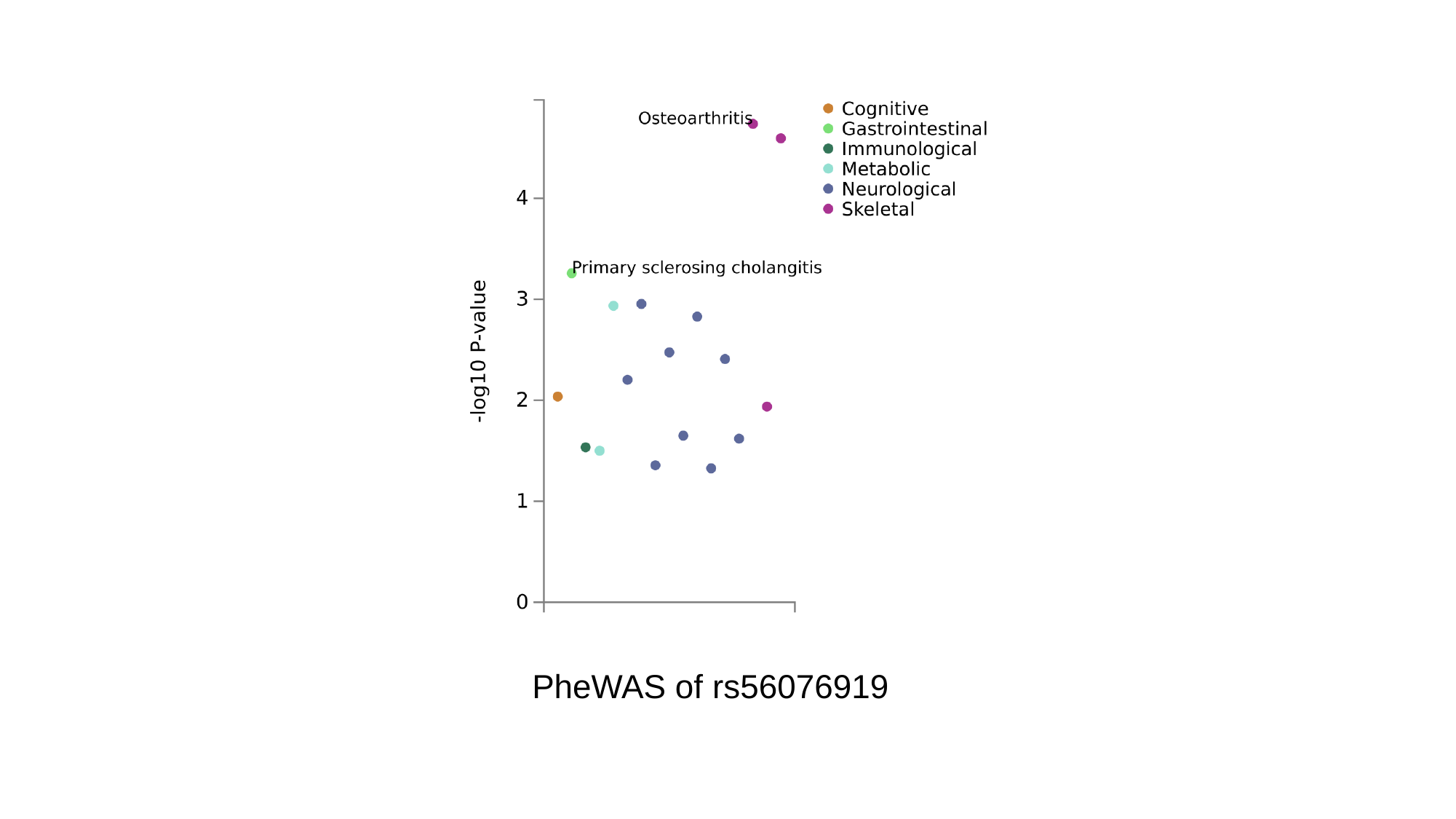

PheWAS of rs56076919

### Slide 4
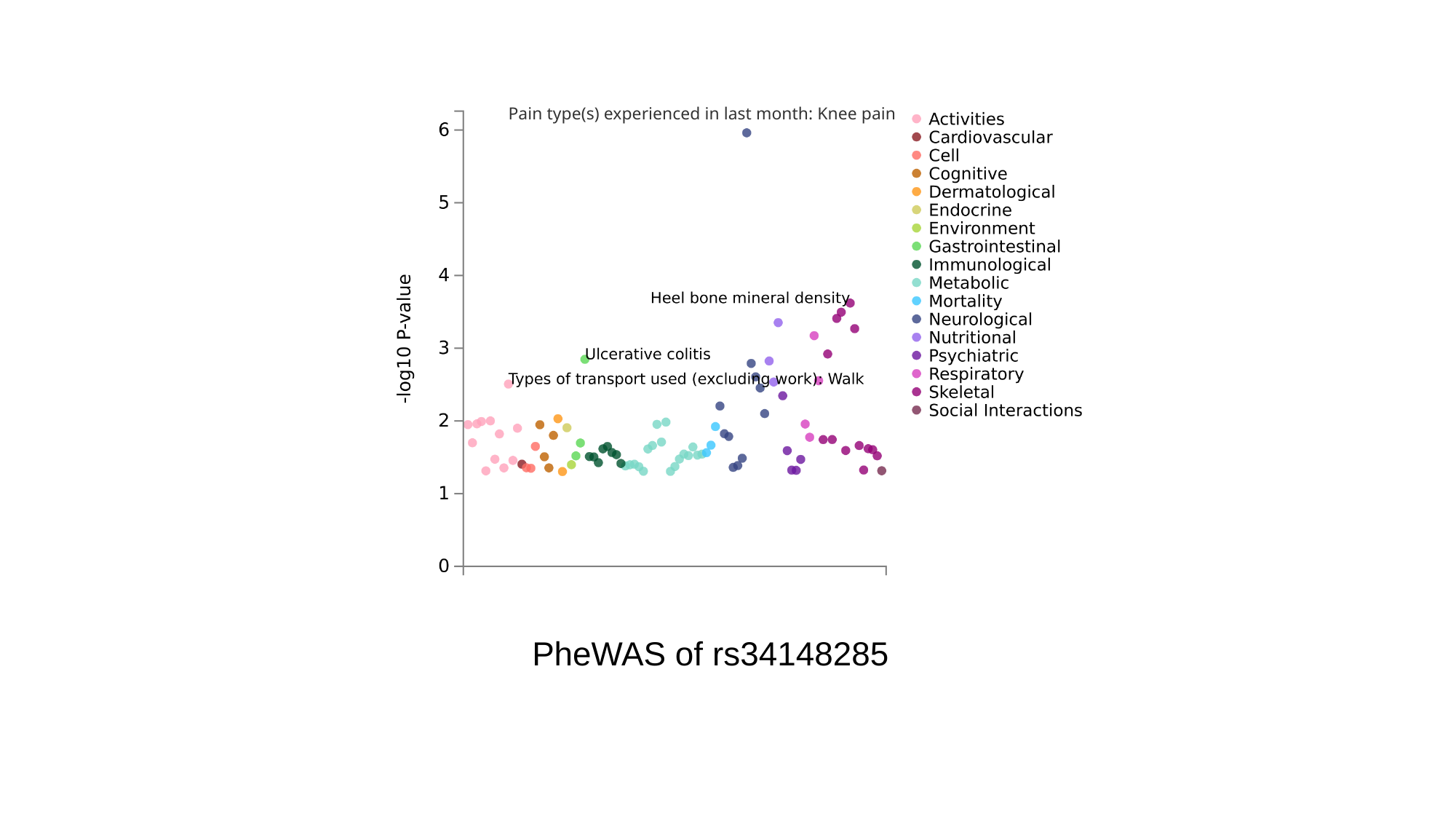

Pain type(s) experienced in last month: Knee pain
PheWAS of rs34148285

### Slide 5
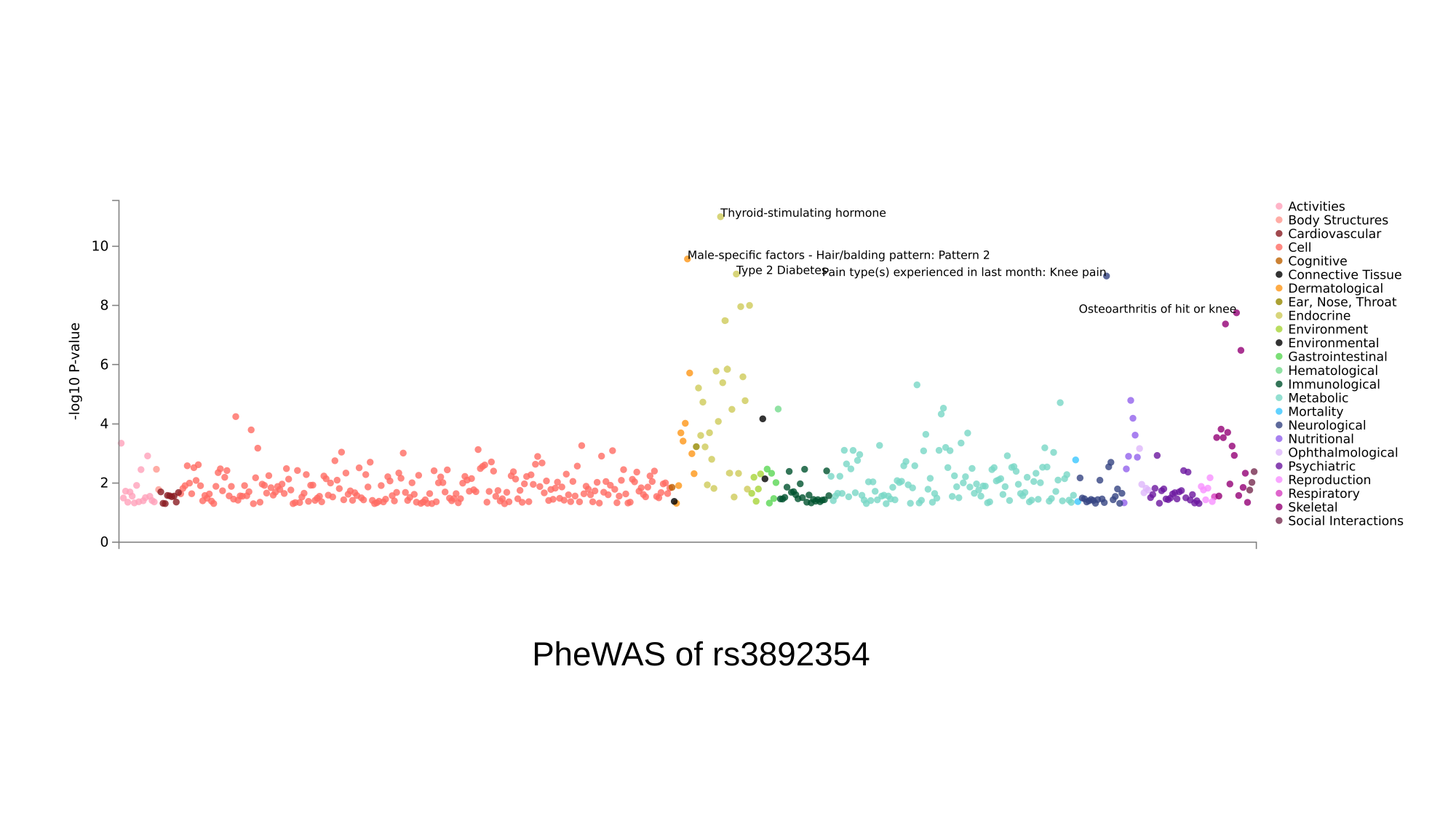

PheWAS of rs3892354

### Slide 6
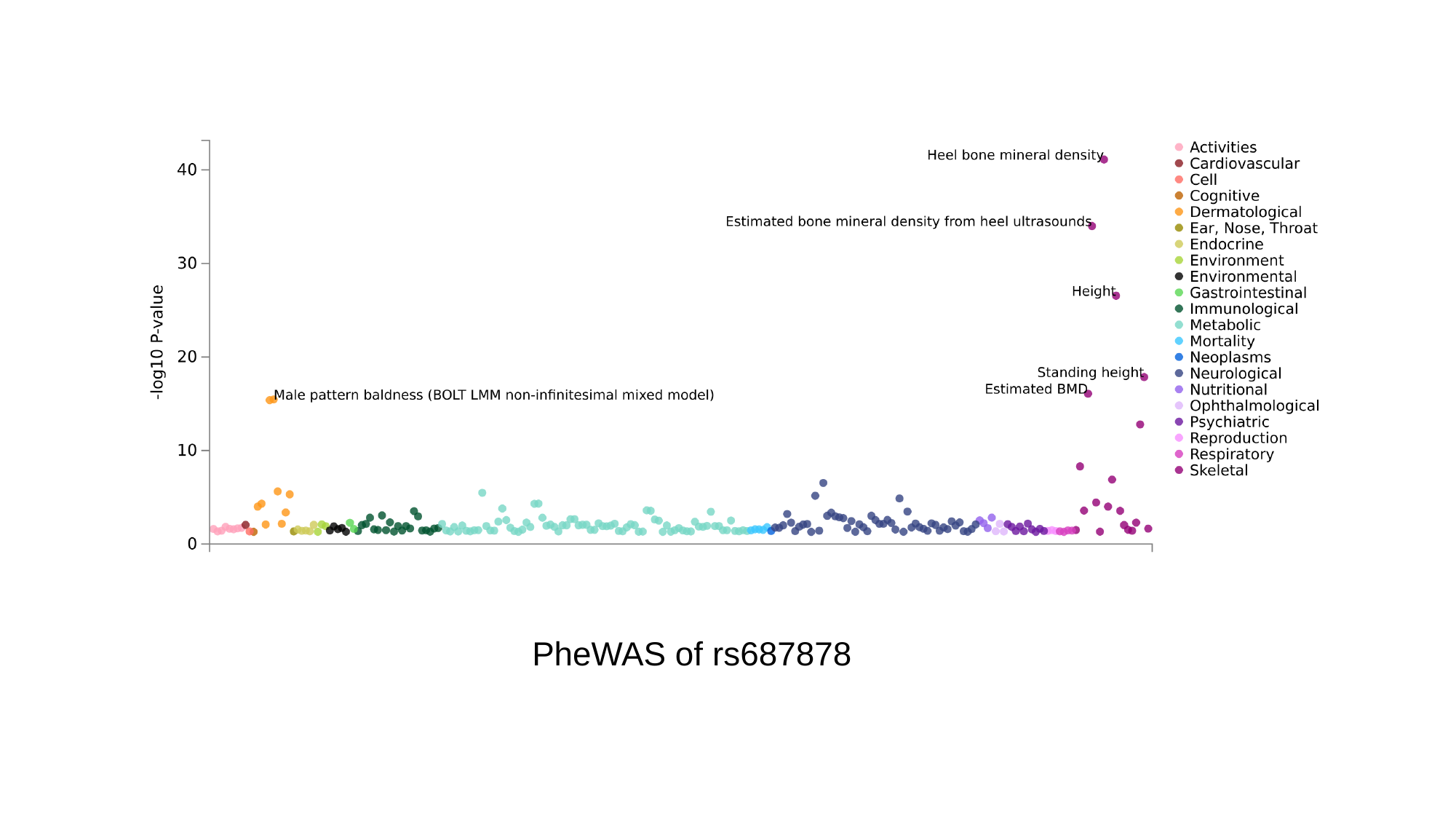

PheWAS of rs687878

### Slide 7
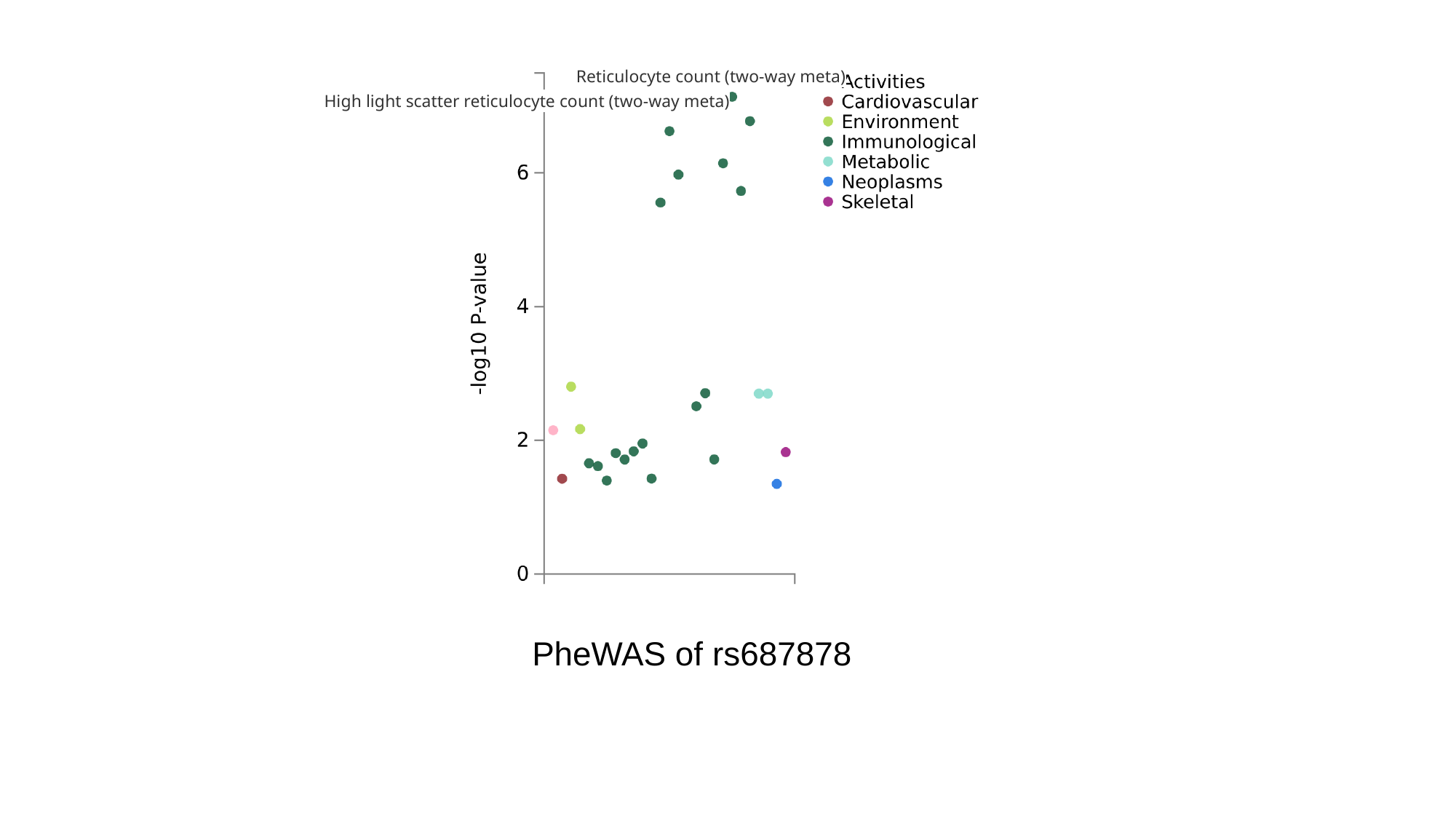

Reticulocyte count (two-way meta)
High light scatter reticulocyte count (two-way meta)
PheWAS of rs687878

### Slide 8
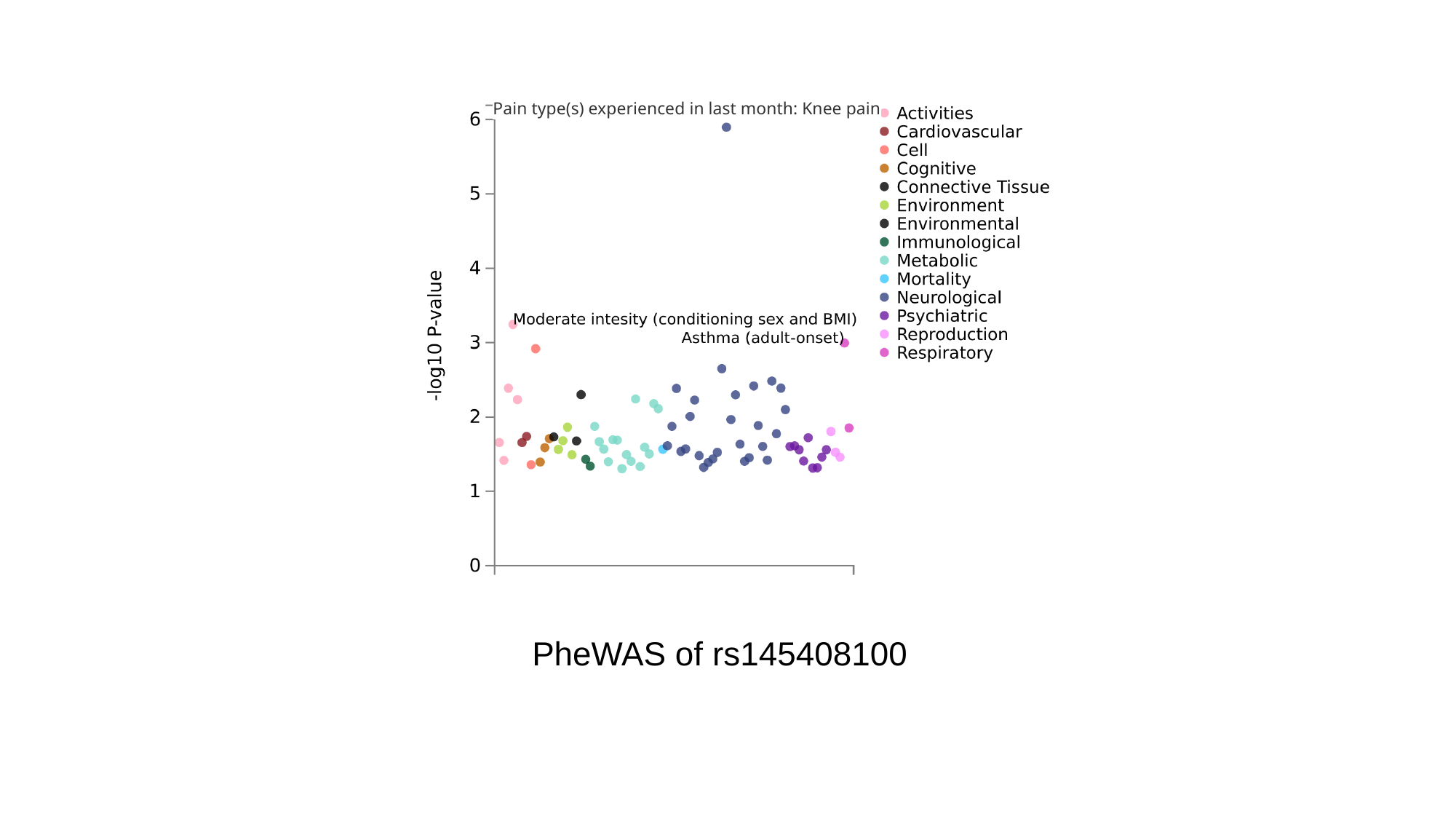

Pain type(s) experienced in last month: Knee pain
PheWAS of rs145408100
