## Supplementary Figures 7 for "A Genome-wide Association Study Identifies Novel Genetic Variants Associated with Knee Pain in the UK Biobank (N = 441,757)"

### Slide 1
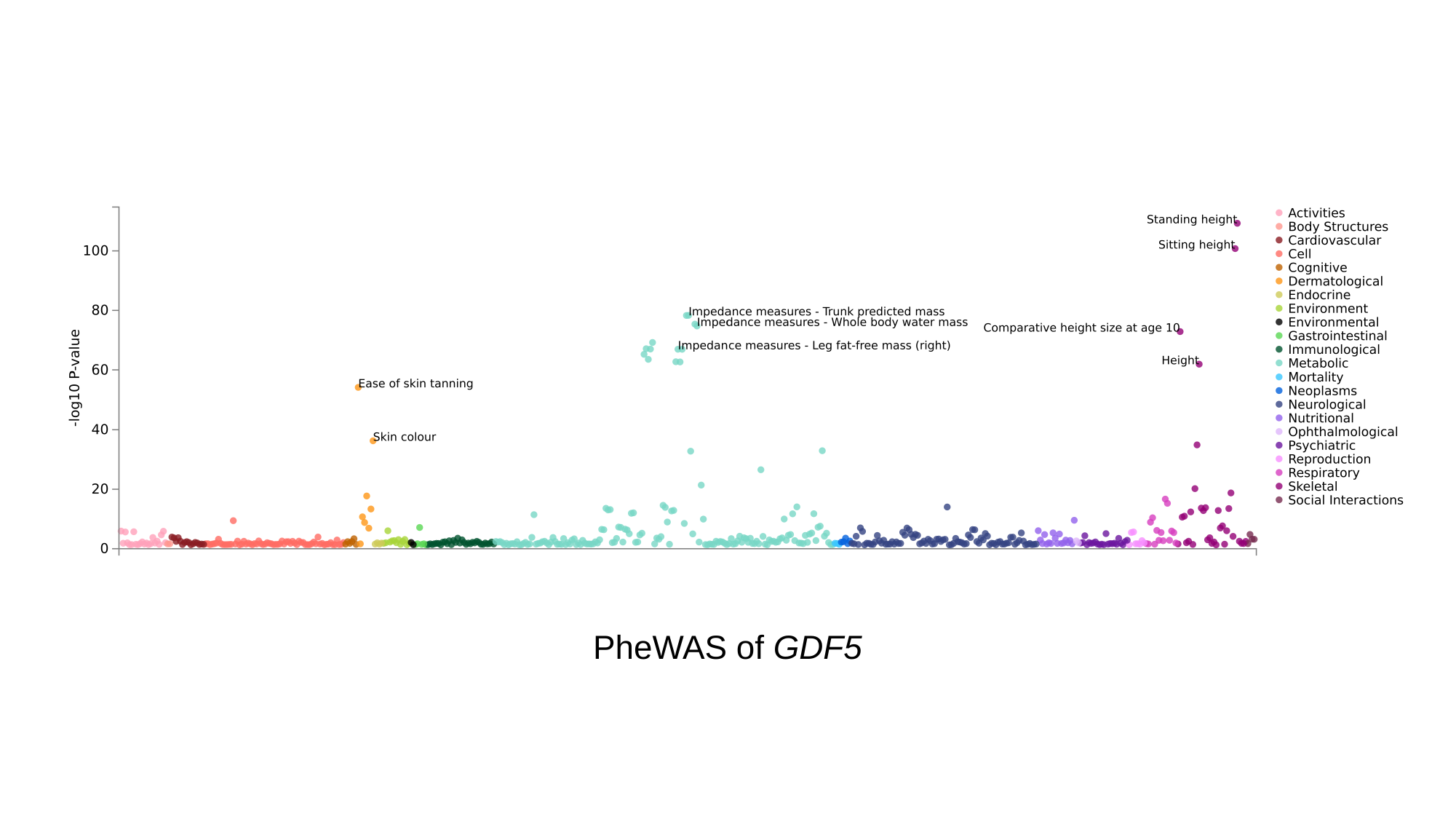

PheWAS of GDF5

### Slide 2
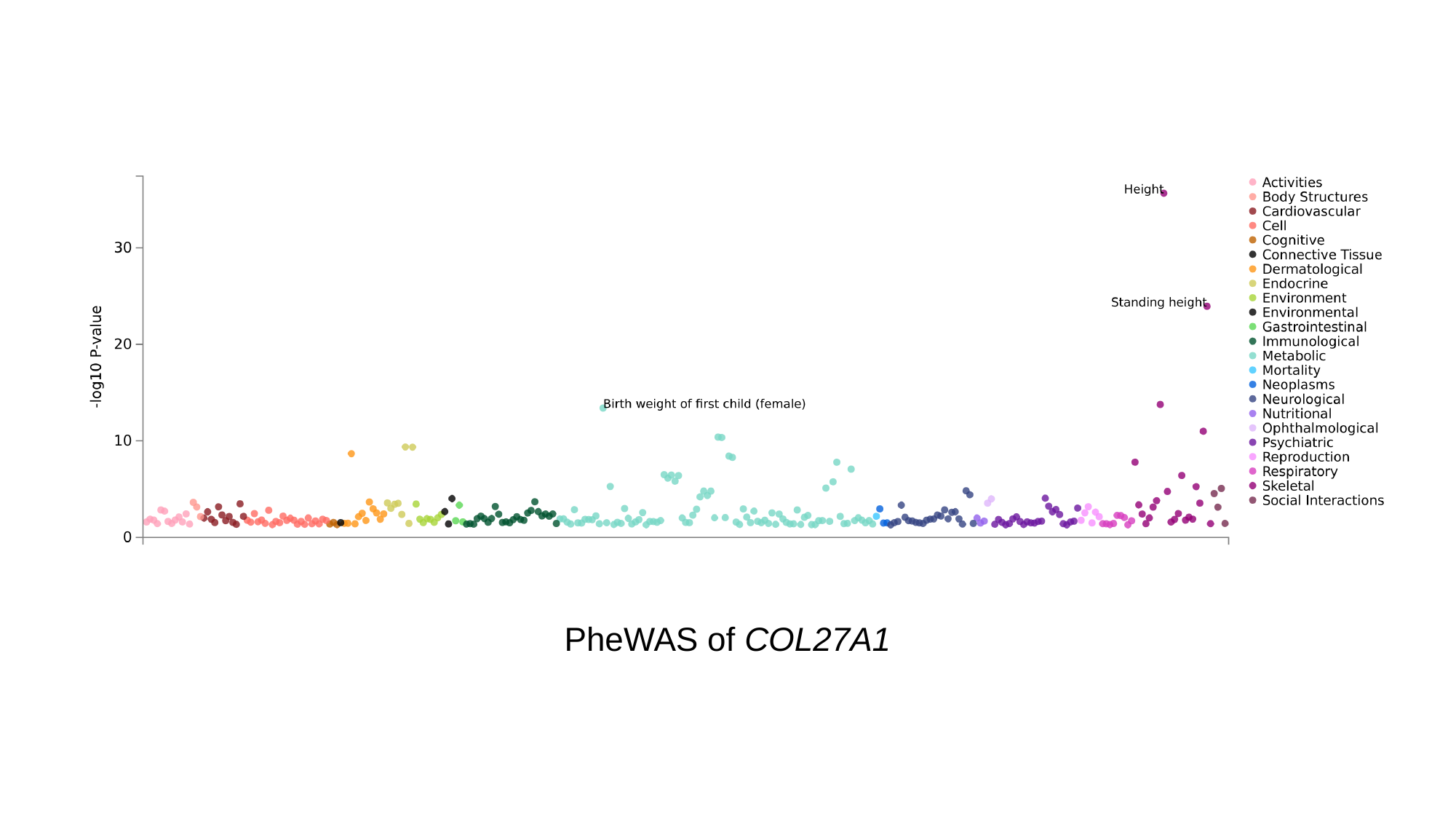

PheWAS of COL27A1

### Slide 3
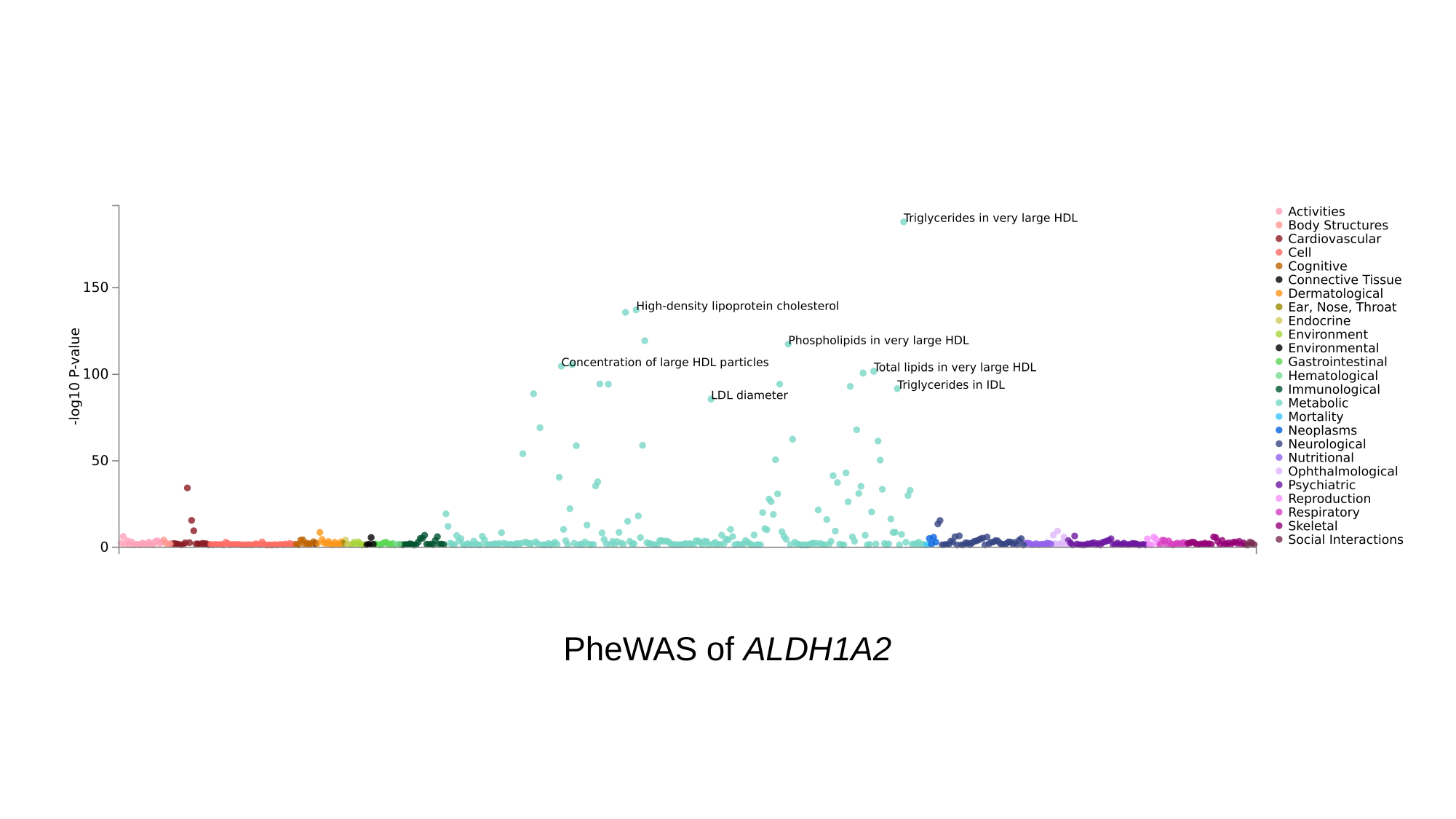

PheWAS of ALDH1A2

### Slide 4
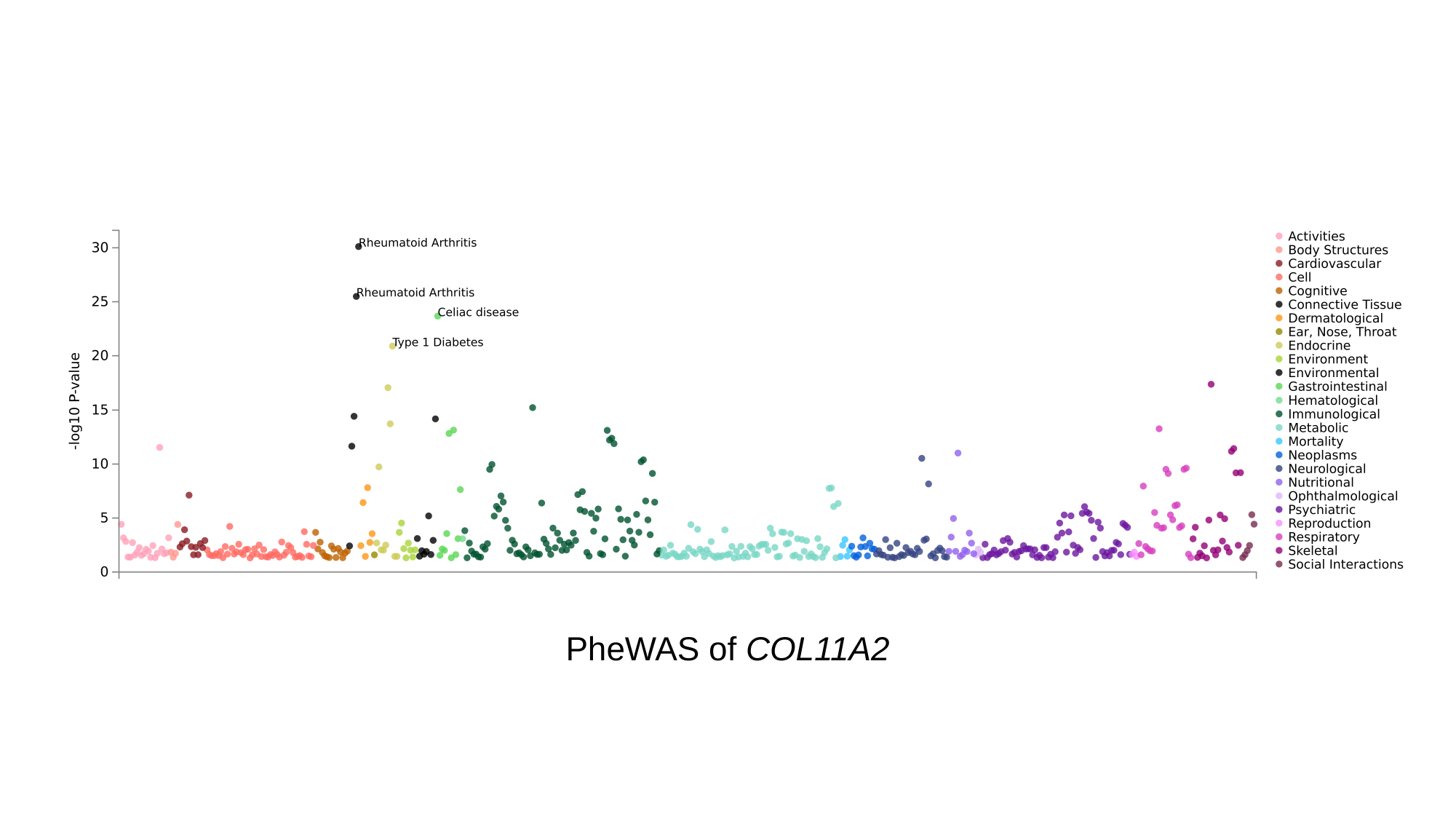

PheWAS of COL11A2

### Slide 5
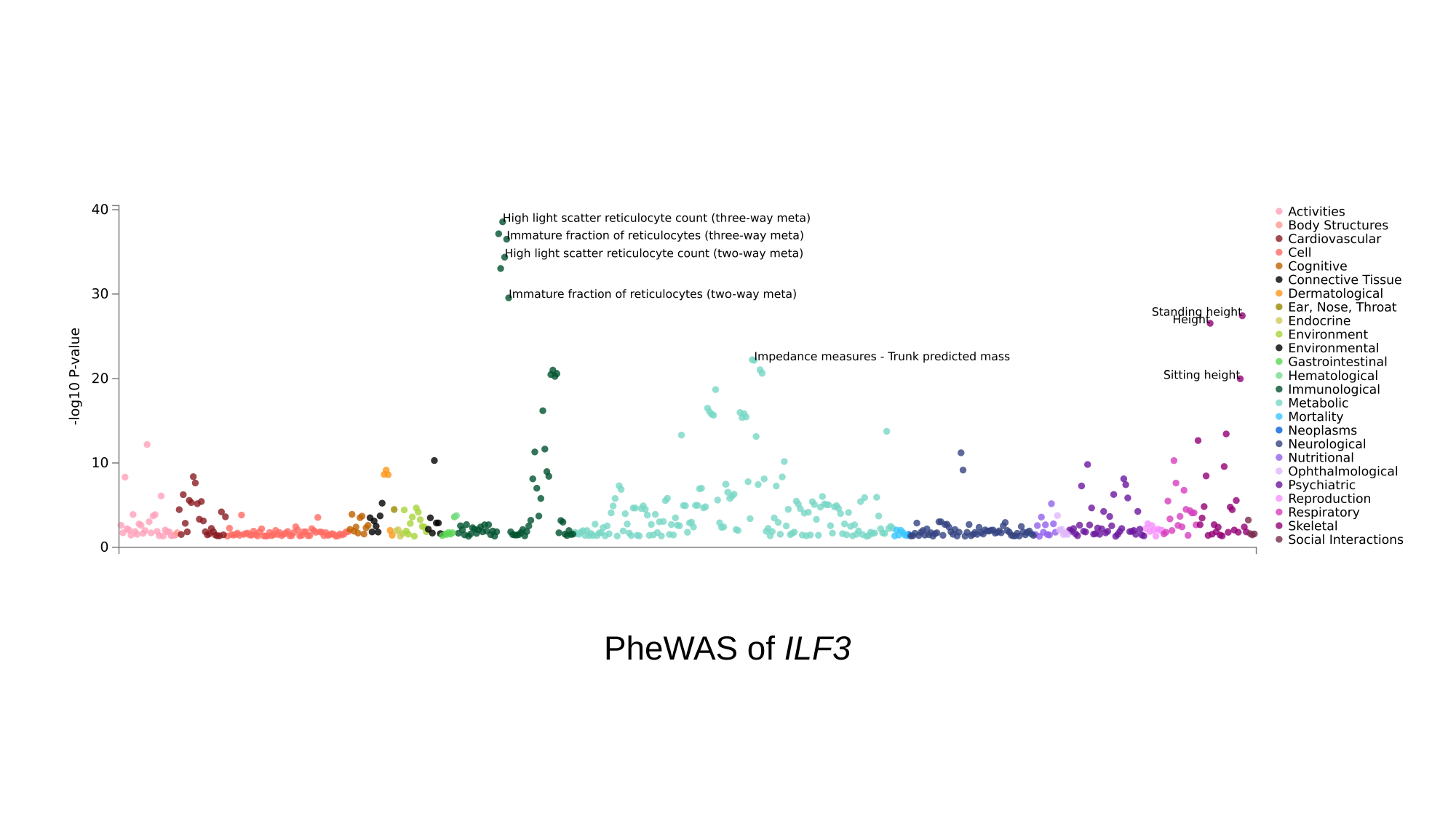

PheWAS of ILF3

### Slide 6
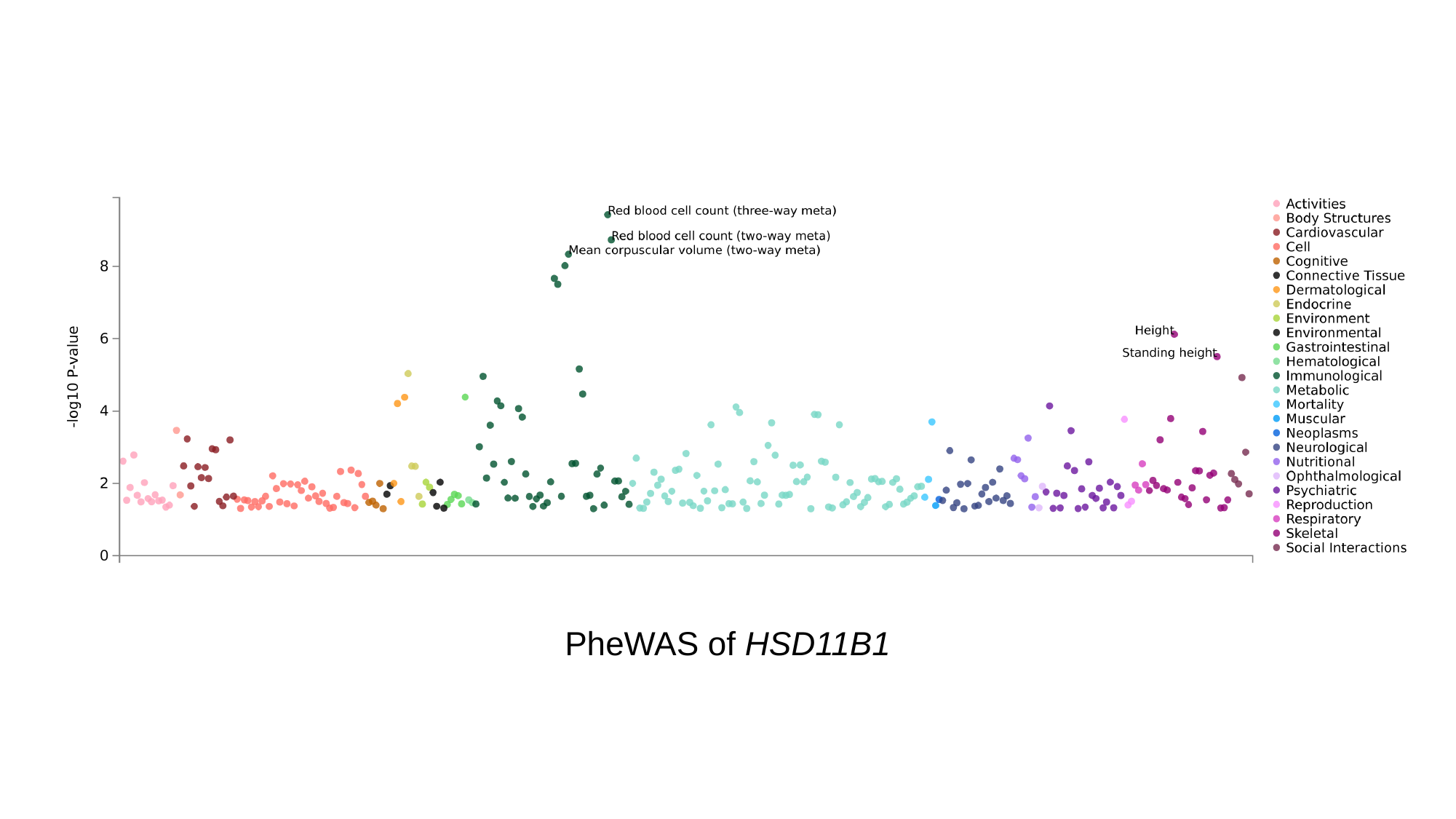

PheWAS of HSD11B1

### Slide 7
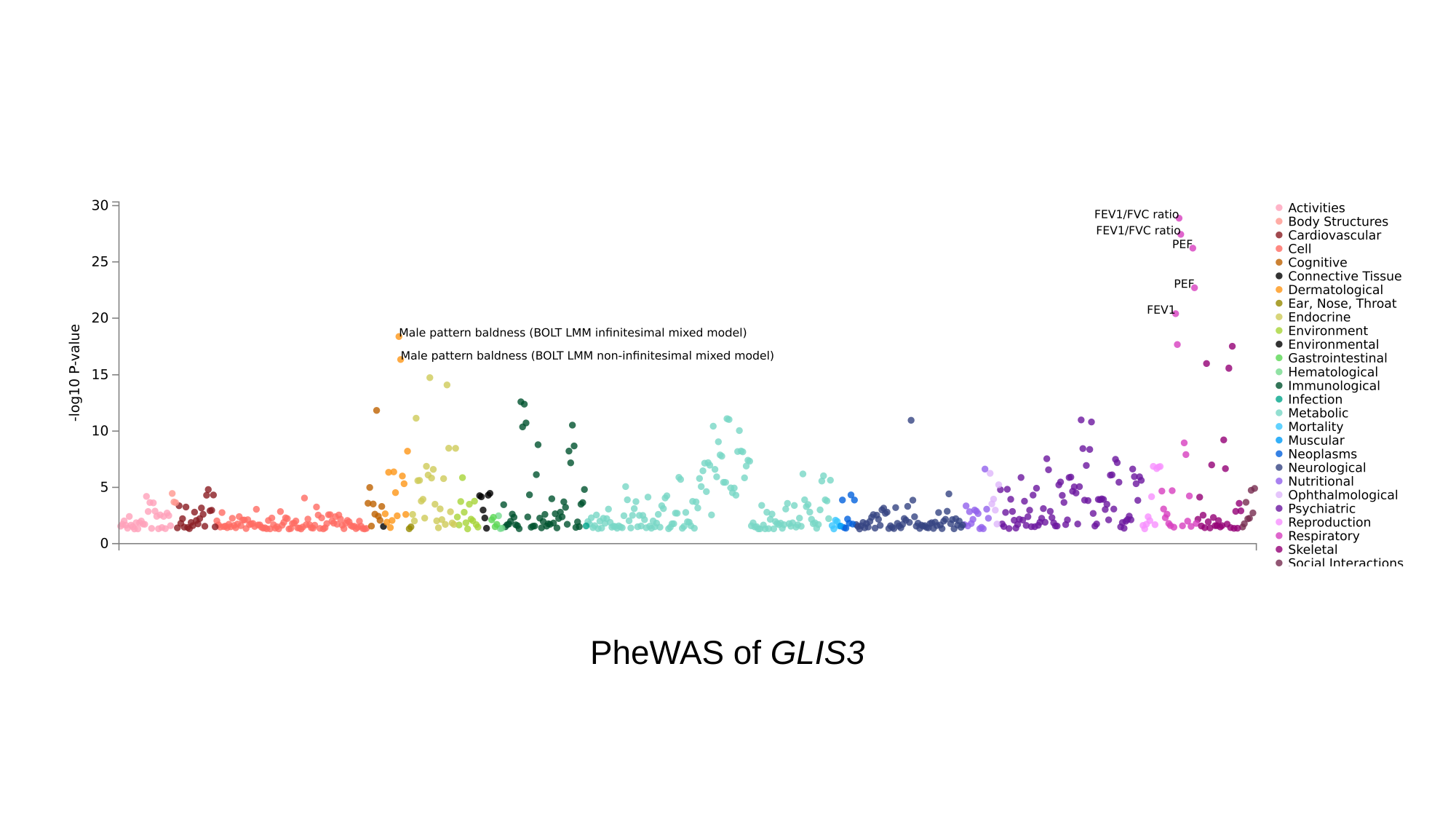

PheWAS of GLIS3

### Slide 8
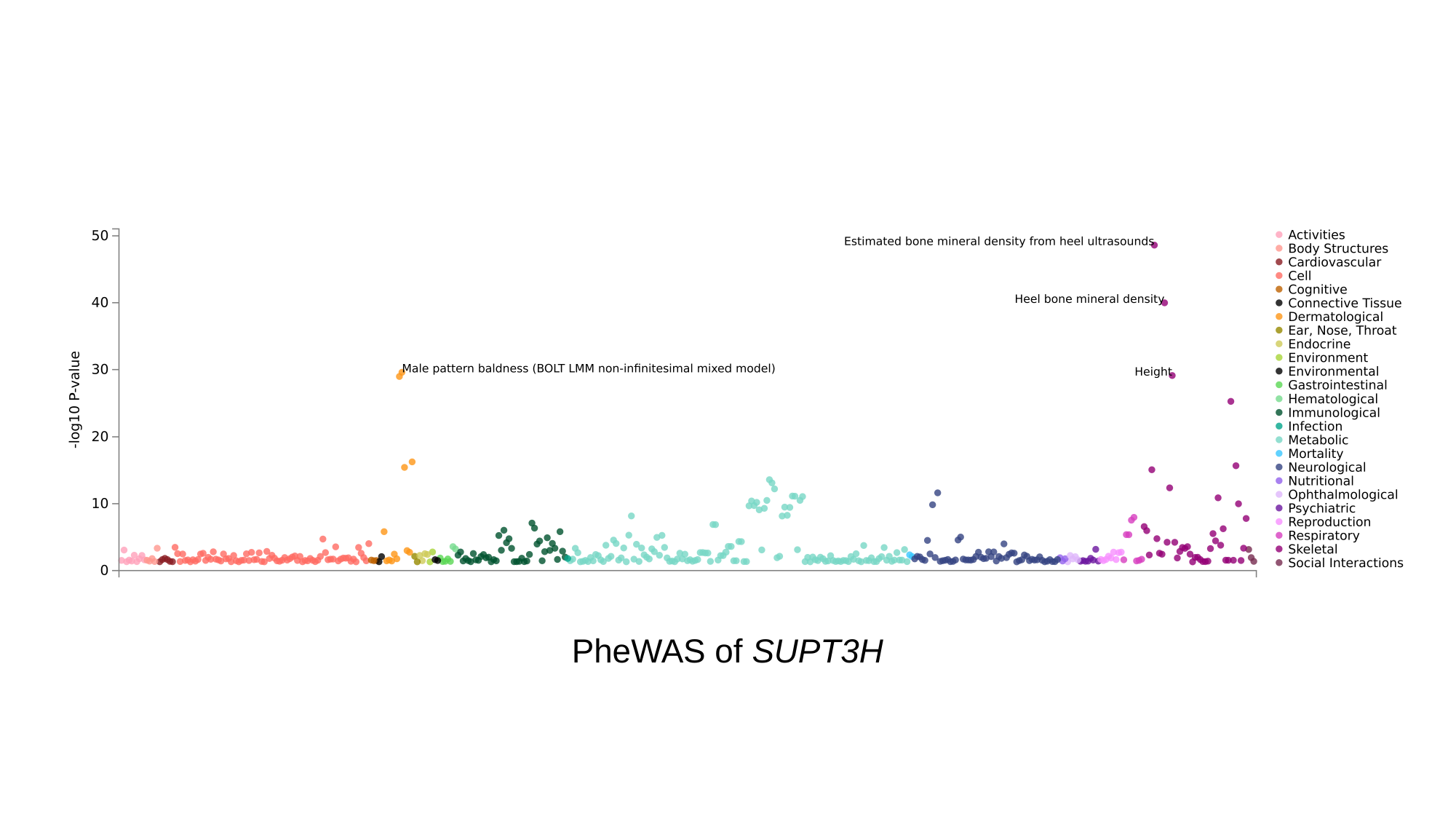

PheWAS of SUPT3H

### Slide 9
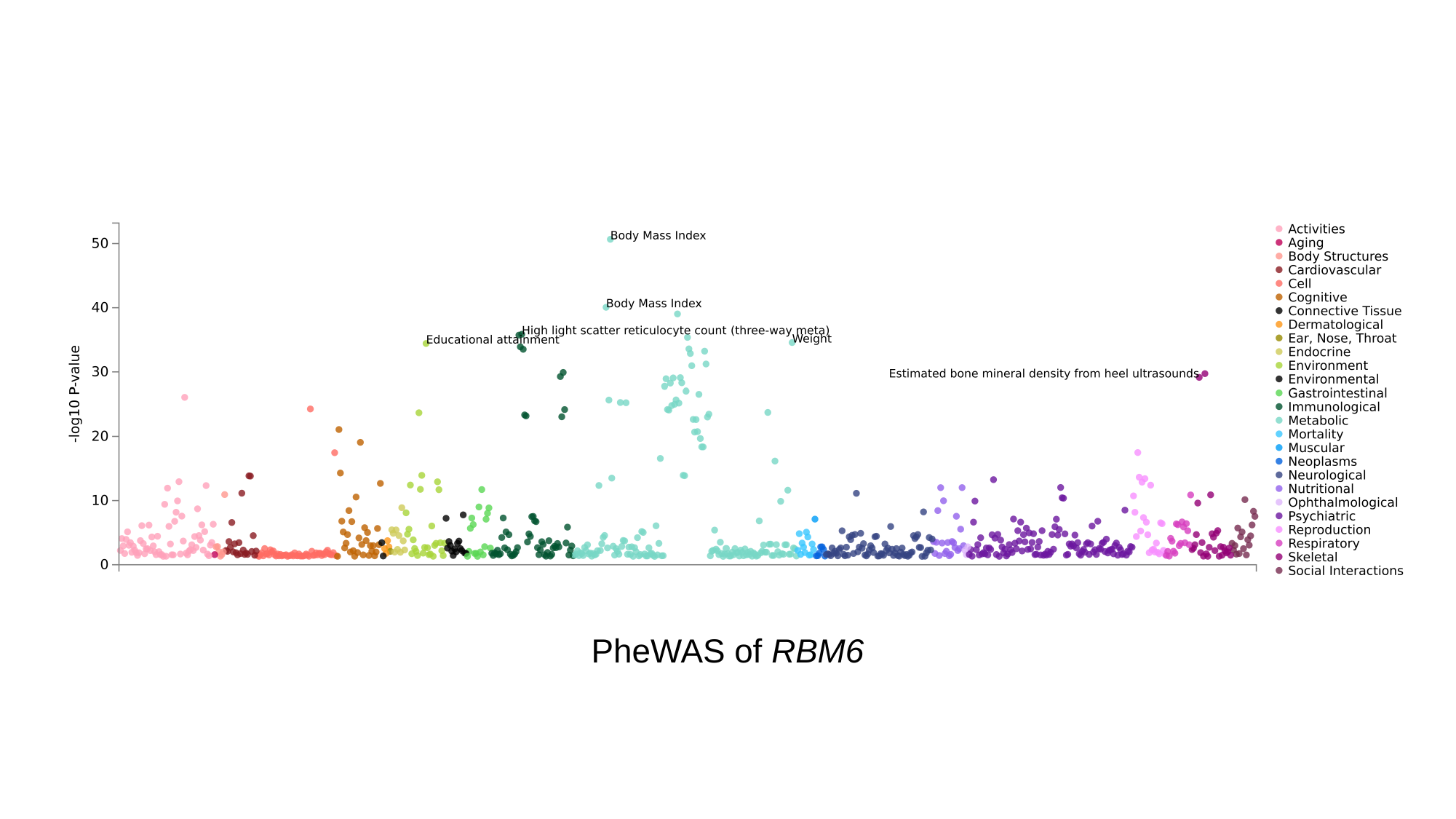

PheWAS of RBM6

### Slide 10
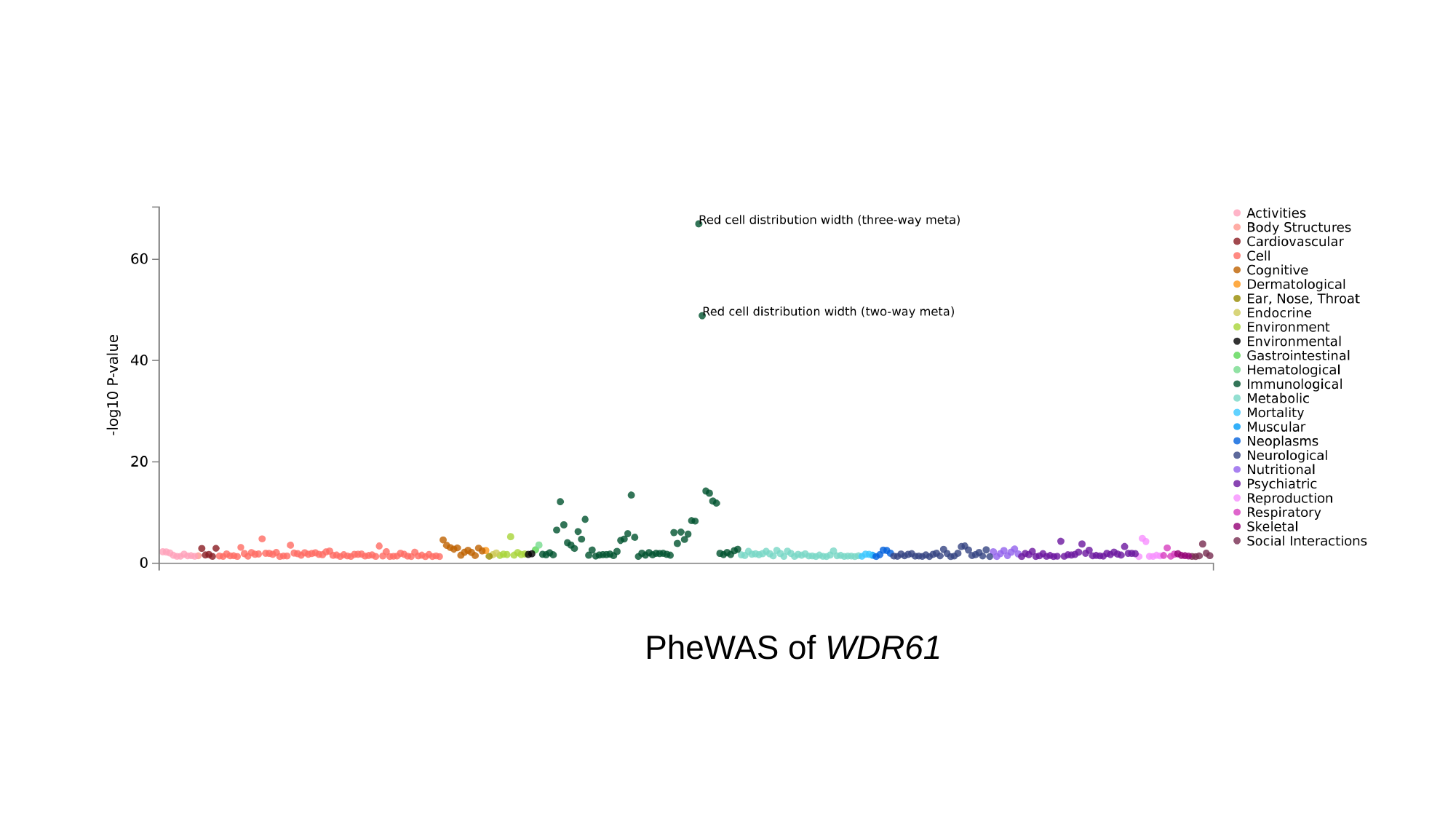

PheWAS of WDR61
