## Supplementary Figures 8 for "A Genome-wide Association Study Identifies Novel Genetic Variants Associated with Knee Pain in the UK Biobank (N = 441,757)"

### Slide 1
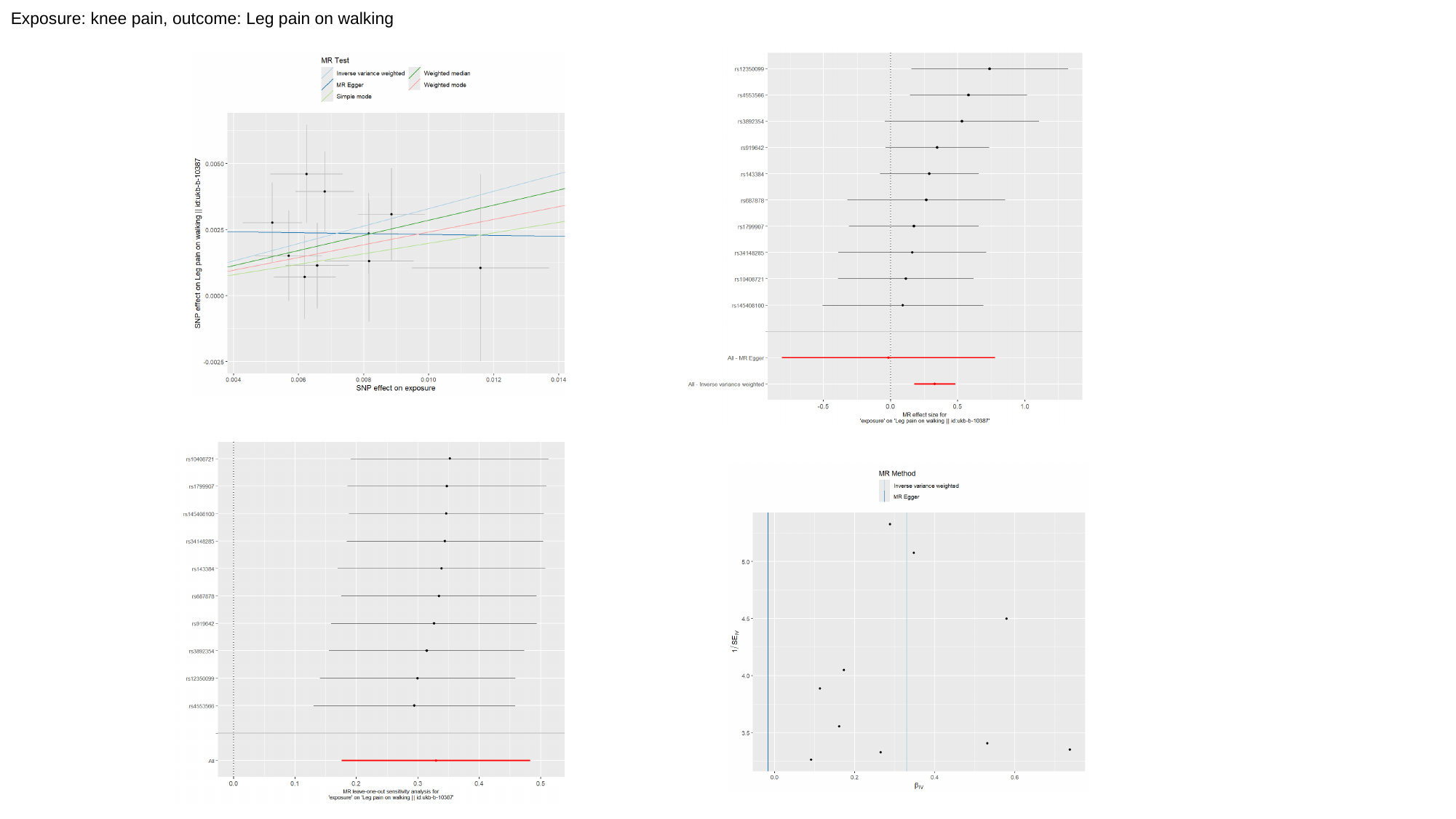

Exposure: knee pain, outcome: Leg pain on walking

### Slide 2
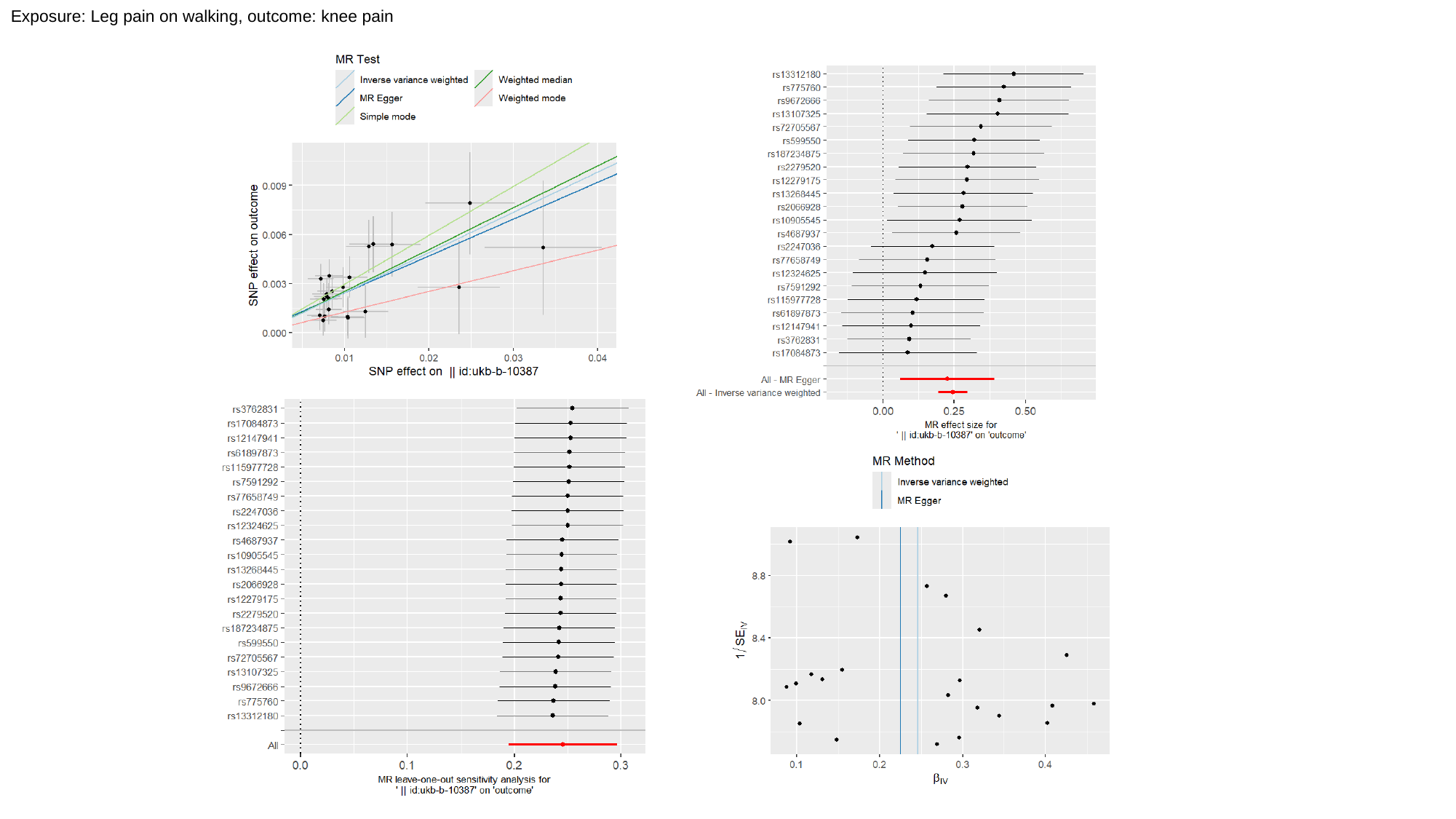

Exposure: Leg pain on walking, outcome: knee pain

### Slide 3
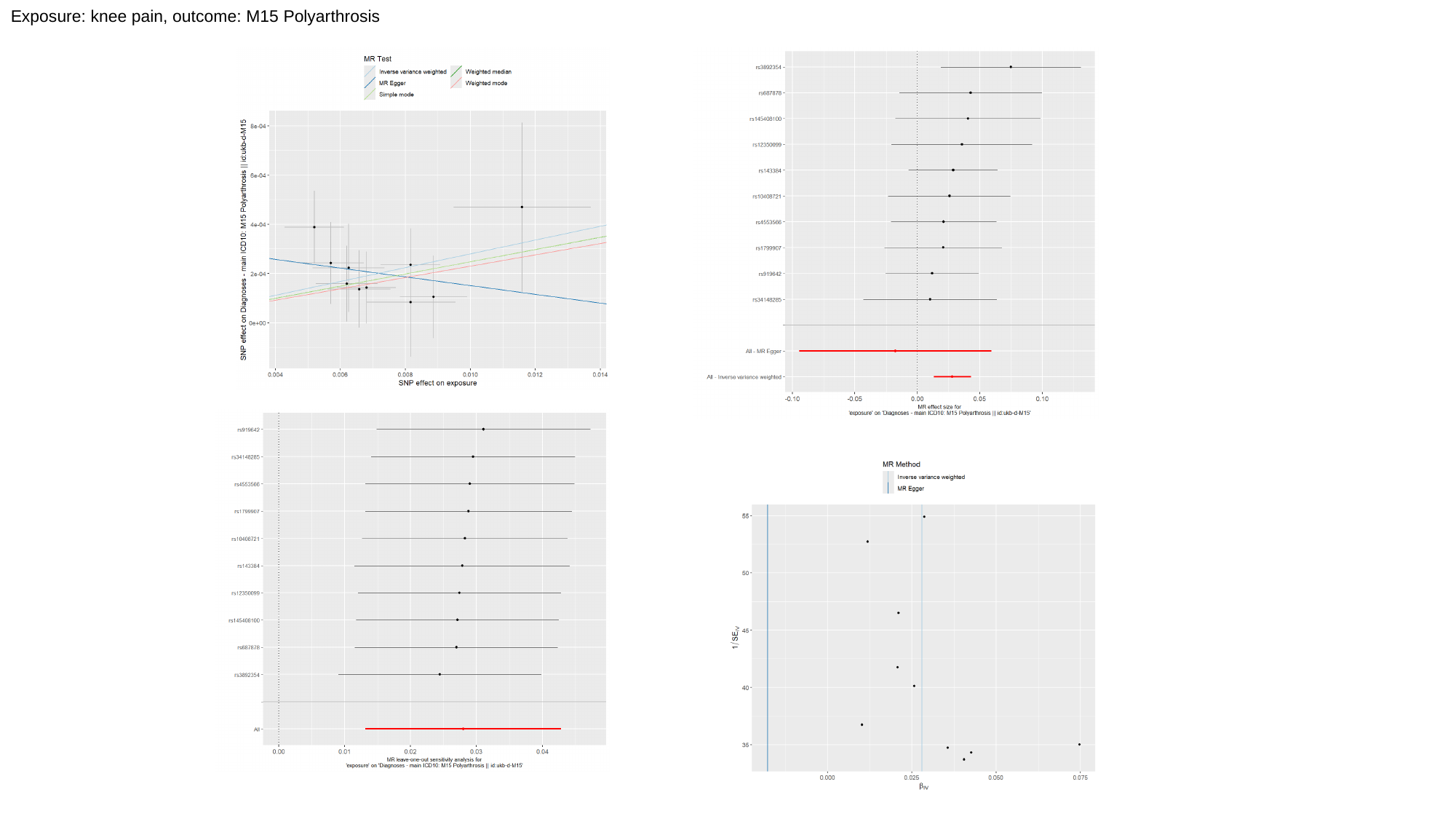

Exposure: knee pain, outcome: M15 Polyarthrosis

### Slide 4
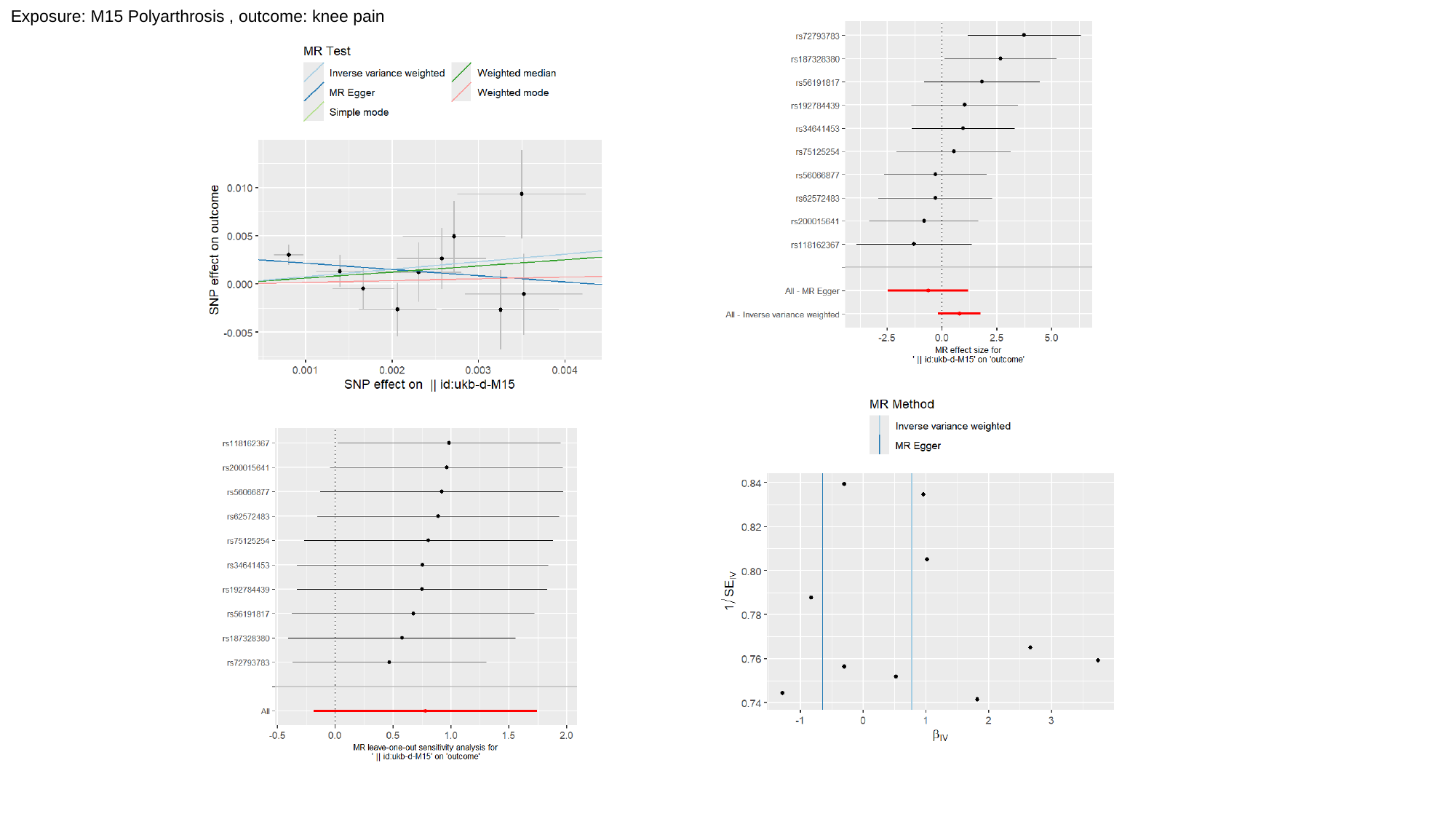

Exposure: M15 Polyarthrosis , outcome: knee pain

### Slide 5
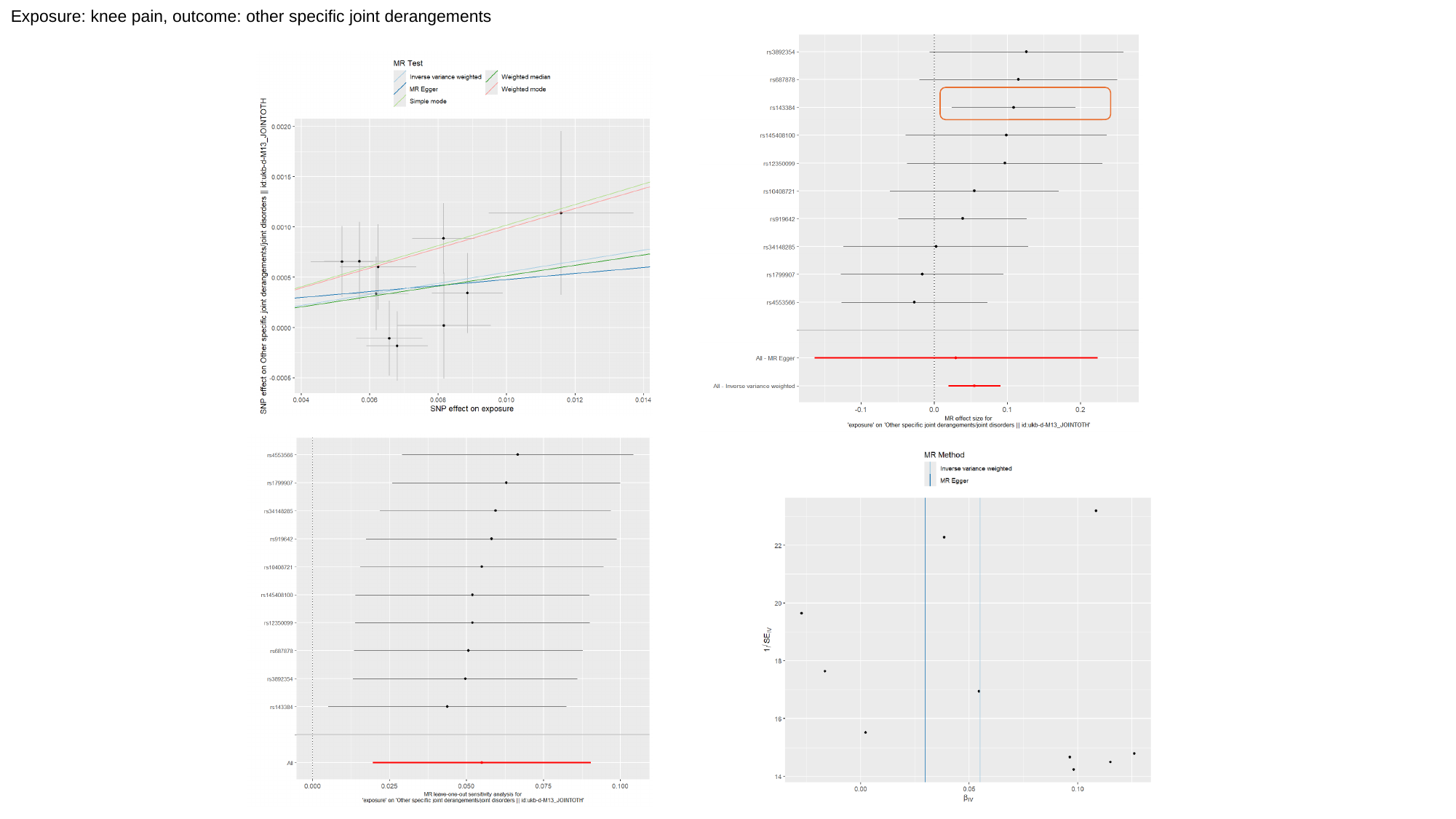

Exposure: knee pain, outcome: other specific joint derangements

### Slide 6
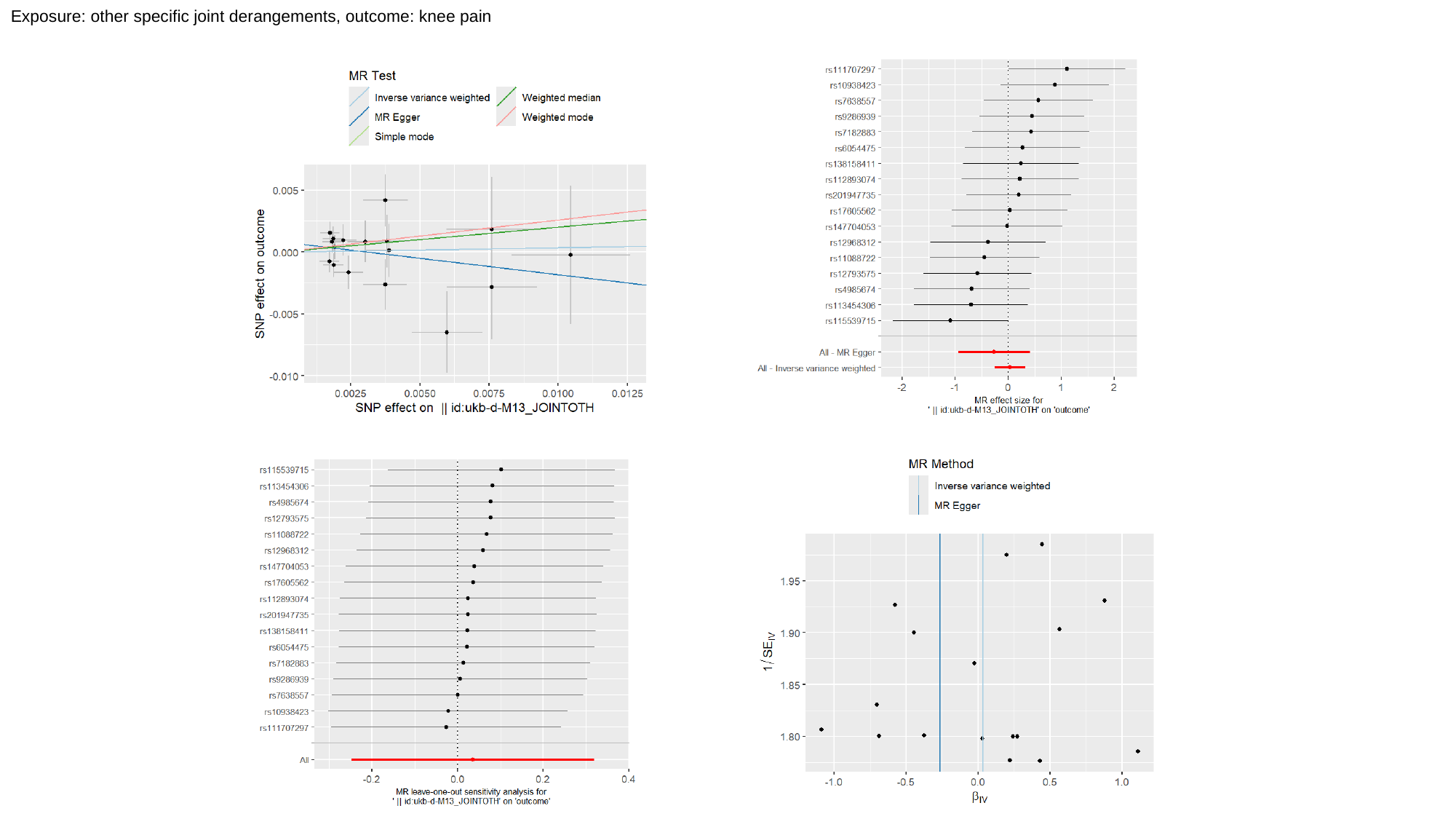

Exposure: other specific joint derangements, outcome: knee pain

### Slide 7
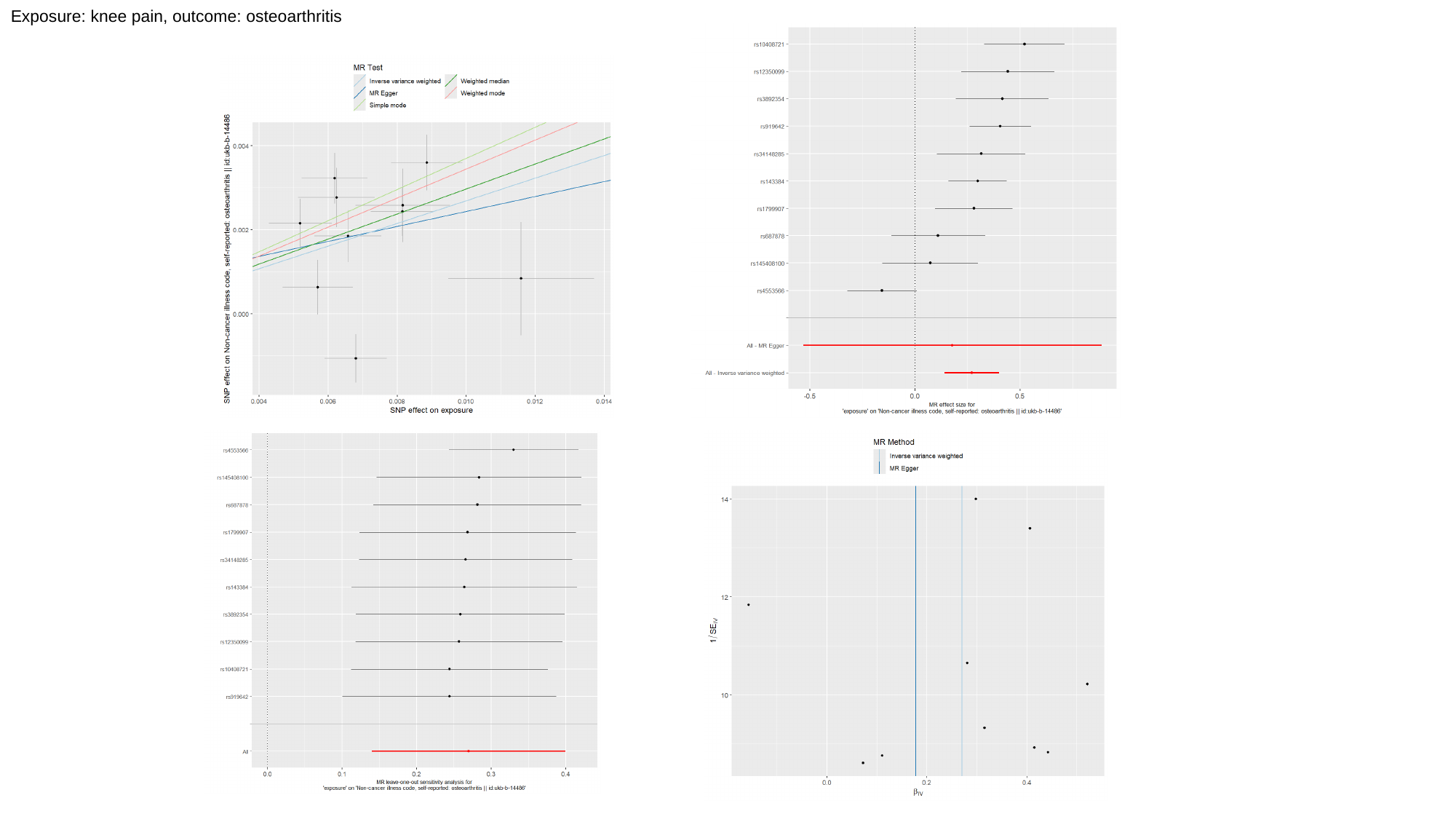

Exposure: knee pain, outcome: osteoarthritis

### Slide 8
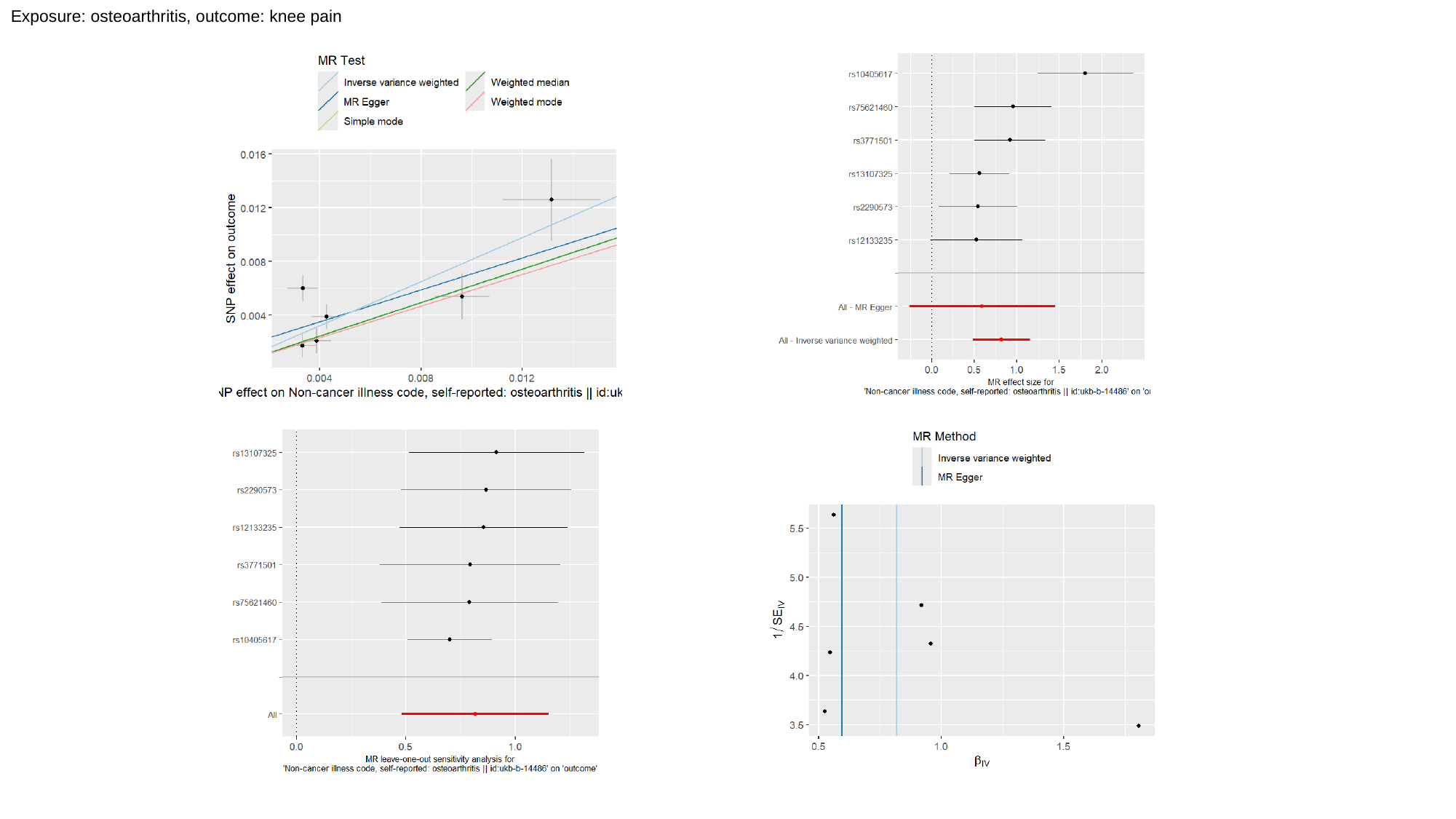

Exposure: osteoarthritis, outcome: knee pain
